# Multi-Ancestry Epigenome-Wide Meta-Analysis Identifies Novel Bulk and Cell-Type-Specific Epigenetic Markers of Asthma with Severe Exacerbations

**DOI:** 10.64898/2026.04.16.26350345

**Authors:** Javier Perez-Garcia, Elena Martin-Gonzalez, Zeyuan Johnson Chen, Mario Martin-Almeida, Jonathan Witonsky, Aditya Gorla, Celeste Eng, Fabian Lorenzo-Diaz, Anne K. Bozack, Jennifer R. Elhawary, Donglei Hu, Scott Huntsman, Ruperto González-Pérez, José M. Hernández-Pérez, Paloma Poza-Guedes, Elena Mederos-Luis, Inmaculada Sánchez-Machín, Jose Rodriguez-Santana, Jesús Villar, Sheryl L Rifas-Shiman, Marie-France Hivert, Emily Oken, Diane R. Gold, Elad Ziv, Elior Rahmani, Esteban G. Burchard, Andres Cardenas, Maria Pino-Yanes

## Abstract

**Background:** Extreme-phenotype comparisons allowed the discovery of novel asthma genetic risk loci. However, this approach remains unexplored in epigenome-wide association studies (EWAS). We aimed to identify bulk and cell-specific methylation markers of asthma with severe exacerbations across diverse ancestry groups.

**Methods:** We conducted a meta-EWAS of 739,543 CpGs in whole blood among 1,192 African American and Latino pediatric populations, comparing non-asthmatics and asthma exacerbators. Genome-wide CpGs were followed up for replication in a meta-analysis across 1,516 ethnically diverse participants and in a cross-tissue evaluation of 393 nasal samples. We conducted differentially methylated region (DMRs), cell-type-deconvoluted, and quantitative trait loci analyses (whole-genome sequencing n=1,668; RNA-seq n=1,209). We examined enrichment in traits, pathways, and druggable genes, and analyzed DNAm predictors of plasma proteins and aging.

**Results:** DNAm at 505 CpGs and 119 DMRs in whole blood were associated with asthma exacerbations (*p*<9x10^-8^, λ=1.05). We replicated 25 CpGs in blood cells, cross-validated 7 in nasal samples, and detected 42 cell-specific DNAm markers mainly driven by T cells. DNAm at 134 CpGs was associated with gene expression in whole blood, including 118 associations with T-cell receptor genes, and 446 CpGs were regulated by ≥1 genetic variant. We found enrichment for previous associations with environmental exposures, immune disorders, immune and inflammatory pathways, and druggable genes by developmental drugs. 21 methylation-predicted plasma proteins, involved in host defense, and one lung aging clock were associated with asthma exacerbations.

**Conclusions:** The first meta-EWAS of extreme asthma phenotypes identified hundreds of novel DNAm markers, suggesting novel methylation biomarkers and candidate drugs for asthma and supporting the role of T cells.

## INTRODUCTION

Asthma is a complex respiratory disease characterized by chronic airway inflammation, reversible expiratory airflow limitation, and a history of respiratory symptoms (1). Asthma symptoms result from inflammatory pathogenic mechanisms, including airway remodeling, epithelial barrier disruption, airway hyperresponsiveness, bronchoconstriction, and mucus hypersecretion (1). This heterogeneous disease is characterized by clinical features (e.g., atopy, immunoglobulin E [IgE] levels, blood and sputum eosinophils, lung function, and fractional exhaled nitric oxide) and endotypes, defined by the type of airway inflammation (T2-high *vs.* T2-low) (2,3).

Asthma remains a major global biomedical concern (2). In 2021, 260 million individuals of all ages had asthma worldwide, which is expected to increase to 275 million by 2050, and >400,000 annual deaths were related to asthma (4). A major contributor to asthma burden are asthma exacerbations, which in their severe form involve life-threatening worsening episodes requiring oral corticosteroid (OCS) treatment and urgent actions, such as hospitalizations or emergency room visits (1,5). These are associated with substantial socioeconomic impact, increasing >$5,000 of annual asthma-related healthcare costs per patient (6).

Asthma results from the interaction between genetic factors and environmental exposures (e.g., smoking, air pollution, allergens, and microbes) (2,3), and the study of epigenetics might bridge the gap between gene-environment interactions and asthma and exacerbation risk (3,7). DNA methylation (DNAm) of cytosines located in 5′-cytosine-phosphate-guanine-3′ dinucleotides (CpG) is the most studied epigenetic modification in asthma (3,7–9). Most epigenome-wide association studies (EWAS) of asthma have focused on whole blood, which may provide information about systemic immune and inflammatory mechanisms, and on nasal tissue, as a non-invasive proxy for the pathophysiology in the lower airways (3,7–10). Most EWAS signals for asthma in these tissues are located in genes involved in T2 inflammation, immune system regulation, eosinophilic response, and epithelial barrier integrity (8,10).

However, our understanding of DNAm markers implicated in asthma, and especially for exacerbations, remains incomplete. Only 3% of thousands of CpGs associated with asthma by EWAS have been replicated (8,9), and 70% of studies lack participants of diverse ancestry (11), limiting their transferability. While bulk DNAm-based studies are valuable for asthma biomarker development, the use of cell deconvolution techniques in asthma remains unexplored (9,12). Moreover, we hypothesize that, as demonstrated in genomic studies (13,14), comparisons of extreme phenotypes (non-asthmatic controls *vs.* asthma with severe exacerbations) could reveal new clinically relevant asthma risk epigenetic loci undiscovered so far.

Therefore, we aimed to identify bulk and cell-specific DNAm markers and predicted plasma protein and epigenetic clocks associated with asthma with severe exacerbations in a meta-analysis across diverse ancestry populations. We performed a meta-EWAS across 2,708 ethnically diverse participants with whole-blood DNAm (n_Discovery_=1,192, n_Replication_= 1,516), a leukocyte-cell-type deconvoluted meta-analysis, and a cross-tissue validation in nasal samples (n=393). We integrated DNAm with whole-blood RNA-seq and whole-genome sequencing (WGS) data and examined underlying pathways, druggable genes, and DNAm predictors of plasma proteins and aging.

## METHODS

### Study Populations

We followed a discovery, replication, and cross-tissue evaluation study design to identify DNAm markers associated with asthma with severe exacerbations. A flowchart for sample selection is described in **Figure S1**. In the discovery phase, we meta-analyzed whole-blood samples from 1,192 admixed and minoritized children and young adults, including 502 African Americans, 466 Puerto Ricans, and 224 Mexican Americans. In the replication phase, we meta-analyzed whole-blood samples from 1,516 participants, including 820 children and adolescents from diverse ancestry, 453 European adults, and 243 Hispanic/Latino children and young adults. We conducted a cross-tissue evaluation in nasal swabs from 393 ethnically diverse adolescents. A brief description of all cohorts is provided in **Table S1**.

The respective ethics committees approved all the studies analyzed in this work, and these followed the Code of Ethics of the World Medical Association (Declaration of Helsinki). All participants provided their signed written consent to participate in each study. For participants aged <18, parents and participants provided signed written consent and assent, respectively. Demographic and clinical data were recorded from all participants through interviews by research staff using standardized questionnaires for each cohort. Characteristics of analyzed studies are described in the **Supplementary Methods**. Briefly:

#### Genes-environments & Admixture in Latino Americans study (GALA II)

Case-control study of pediatric asthma among Latino children from the United States (US) (15). Children and young adults (aged 8-21 years old) who self-identified as Hispanic/Latino with four Hispanic/Latino grandparents were recruited between 2006 and 2014 through clinical and community-based centers in different regions of US and Puerto Rico. GALA II was approved by the Human Research Protection Program (HRPP) Institutional Review Board (IRB) at the University of California, San Francisco (UCSF) (UCSF-IRB No. 10-00889). We analyzed whole-blood samples from 466 Puerto Ricans and 224 Mexican Americans in the discovery phase, and 84 Puerto Ricans and 159 Mexican Americans in the replication phase.

#### Study of African Americans, Asthma, Genes & Environments (SAGE)

Case-control study of pediatric asthma conducted in parallel with GALA II, except that participants self-identified as African American and reported four African American grandparents (15). SAGE was approved by the HRPP-IRB of UCSF (UCSF-IRB No. 10-02877). We analyzed whole-blood samples from 502 African Americans in the discovery phase.

#### Project Viva

Pre-birth cohort study of mothers and their offspring recruited between 1999 and 2002 during the first prenatal visit at a multispecialty medical group in Massachusetts (US) (16,17). The Institutional Review Board of Harvard Pilgrim Health Care reviewed and approved all study protocols. Mothers and their offspring were followed up in in-person interviews during the pregnancy until the present. We analyzed 358 and 462 whole blood samples collected at ages 7 and 13, respectively, in the replication phase, and 393 nasal swabs collected at age 13 in the cross-tissue evaluation stage.

#### Genomics and Metagenomics of Asthma Severity (GEMAS)

Ongoing case-control study aimed at investigating the molecular basis of asthma exacerbations (18). Participants with and without asthma aged 8-85 years were recruited between 2018 and 2024 in several Allergy and/or Pulmonology hospital departments in Spain. GEMAS was approved by the ethics committees of the participating centers (approval 29/17 for hospitals in the Canary Islands). We analyzed 453 whole-blood samples from adults in the replication phase.

### Main outcome

The main outcome was asthma with severe exacerbations. Controls were defined as participants without a medical record of asthma, asthma symptoms, asthma medications, or any other chronic respiratory condition. Cases were defined as participants with asthma reporting severe exacerbations in the past year.

Asthma was defined in GALA II and SAGE by 1) physician diagnosis of asthma, 2) recent use of asthma controller/reliever medication, or 3) occurrence of ≥2 asthma symptoms (cough, wheeze, or shortness of breath) in the two years before enrollment. Asthma was similarly defined in each replication cohort based on physician diagnosis as described in the **Supplementary Methods**. In all studies, severe asthma exacerbations were defined based on the presence of emergency care, hospitalizations, and/or OCS prescription in the past year due to asthma (5).

### Genome-wide DNAm measurement and quality control

In GALA II and SAGE, genomic DNA was extracted from whole-blood EDTA samples using the Wizard Genomic DNA Purification Kits 76 (Promega, Fitchburg, WI). DNAm was measured in 856,154 CpGs across the genome using the Infinium Illumina MethylationEPICv1 BeadChip (Illumina, San Diego, CA) following the manufacturer’s protocols. A new DNAm batch was generated in addition to previously published data (19,20), and a standardized quality control (QC) of DNAm data was conducted using the *ENmix* and *ewastools* R packages, as described in the **Supplementary Material** (21,22).

Briefly, we conducted probe-level and sample-level QC by filtering those with low-quality data and/or high missingness rate. We removed individuals with first- or second-degree relatedness and those potentially cross-sample contaminated. We filtered out potentially problematic probes, including probes with mapping inaccuracies or with manufacturer issues, multimodal distribution, potentially capturing genetic variation rather than DNAm, and/or not annotated to autosomes. The number of samples and CpGs that passed QC is reported in **Table S2**. Beta values were log-transformed to M-values to minimize skewed distributions (23), and all analyses were conducted on M-values.

DNAm matrix was corrected for known batch effects using the ComBat algorithm (*sva* R package) while adjusting for the outcome and covariates of subsequent analyses (24). To capture orthogonal sources of unknown technical variation, we performed a principal component analysis (PCA) on residuals of beta values of negative control probes after regressing for the known technical batch (25). Based on a scree plot of explained variance, two technical PCs were included as covariates in subsequent analyses (**Figure S2**).

### Cell-type deconvolution in blood

We used the semi-supervised Bayesian method BayesCCE to infer blood cell type proportions from DNAm data (26). We used a subset of 236 participants from the GALA II study with available whole-blood DNAm and complete blood count (CBC) data to train a model to impute CBC for remaining participants (neutrophils, lymphocytes, eosinophils, monocytes, and basophils). CBCs were rescaled to a sum proportion of 100%. When included as covariates to control for cell-type heterogeneity, the lowest abundant cell type (i.e., basophils) was excluded to avoid collinearity.

### Statistical analyses

A flowchart of analyses conducted in this study is reported in **Figure 1**.

**Figure 1.**
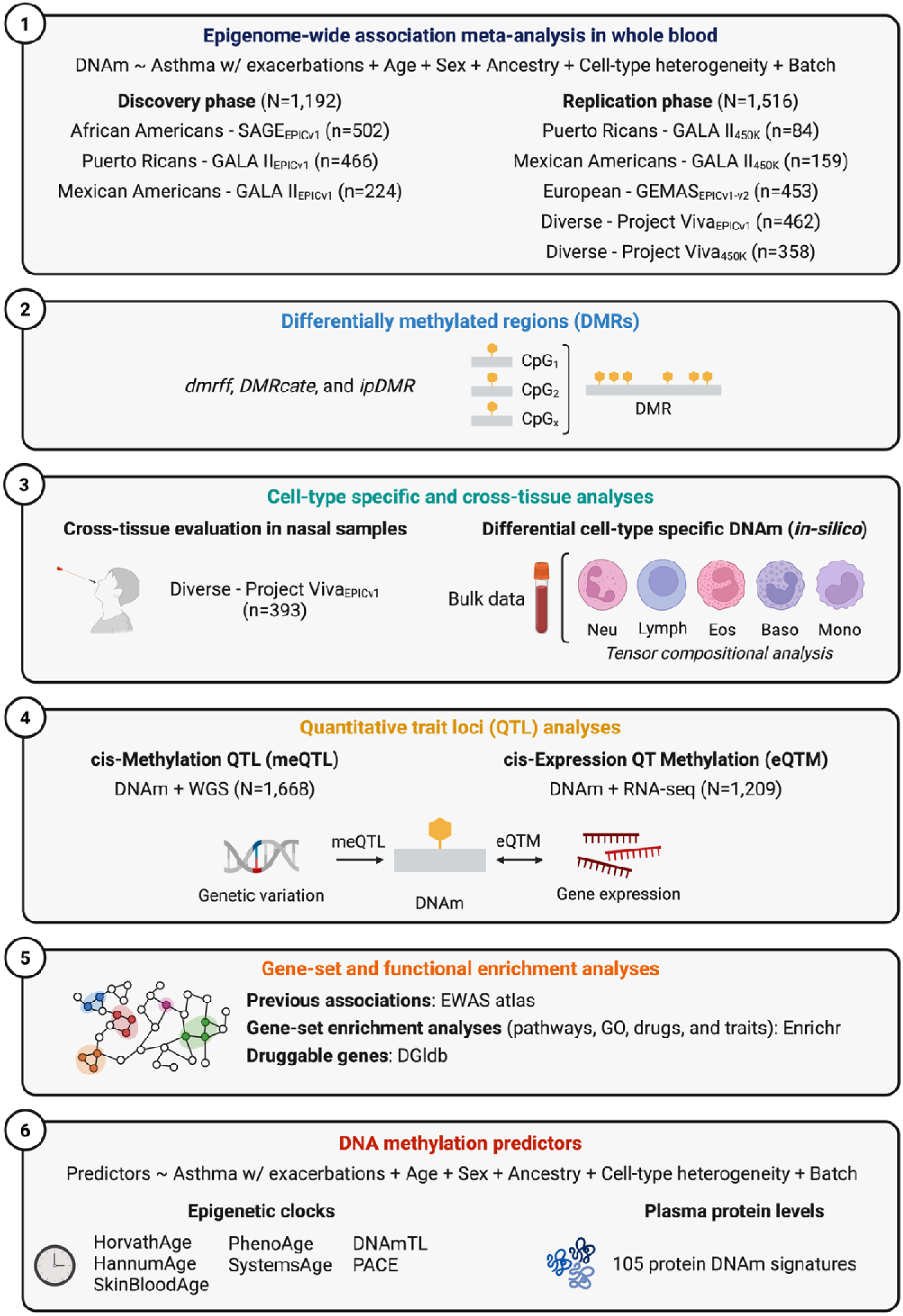
Flowchart of study design. DNAm: DNA methylation, Neu: Neutrophils, Lymph: Lymphocytes, Eos: Eosinophils, Baso: Basophils, Mono: Monocytes, WGS: Whole-genome sequencing, GO: Gene ontologies.

#### Epigenome-wide association study (EWAS)

We tested associations between DNAm levels (M-values) at CpGs across the genome with asthma and exacerbations using robust linear regression models implemented in the *limma* R package (27). Models were adjusted for age, sex, genetic ancestry (the first 2 PCs of WGS data), blood cell counts (predicted percentages of neutrophils, lymphocytes, eosinophils, and monocytes), and technical variation (the first 2 PCs of negative control probes). We conducted analyses in each ethnic group from the discovery phase (African Americans, Puerto Ricans, and Mexican Americans) and then meta-analyzed using METASOFT (739,543 CpGs were available in ≥two ethnic groups) (28). Fixed-effects and random-effects models were applied to CpGs with low (*p*≥0.05) or high (*p*<0.05) heterogeneity, respectively, based on Cochran’s Q test. Given that the actual test-statistic inflation in EWAS can be overestimated by λ_GC_ when many CpGs show significant associations, genomic and bias inflation was examined through Q-Q plots and λ_GC_ and corrected using the BACON Bayesian method (29). Genome-wide significance was declared at *p*<9×10^-8^, an empirical threshold controlling the 5% family-wise error rate for studies using the EPICv1 array (30). Probes were annotated to the human GRCh38/hg38 genome assembly using the Illumina manifest file v1.0 B5, and those without gene annotation were annotated to the nearest gene using *rGREAT* R package (31).

We characterized novel associations by comparing our findings with previous EWAS of asthma using EWAStoolkit at the level of CpGs and genes (*rGREAT* annotations) (31,32). We queried the terms allergic asthma, allergic asthmatic children, asthma remission, asthma rhinitis, asthma, atopic asthma, childhood asthma, and non-atopic asthma. We also searched PubMed for EWAS of asthma exacerbations (33–35).

#### Replication analyses

Genome-wide CpGs identified in the discovery phase were followed for replication in 1,516 participants with available whole-blood DNAm data from the GALA II–450K, GEMAS, and Project Viva studies. DNAm measurements and QC for each dataset are described in the **Supplementary Methods**. Briefly, we analyzed 84 Puerto Ricans and 159 Mexican American children and young adults from GALA II, independent from discovery datasets (Infinium Methylation450K Beadchip), 453 European adults from GEMAS (Infinium MethylationEPICv1 and v2 Beadchips), and 358 mid-childhood ethnically diverse children (age 7, Infinium Methylation450K Beadchip) and 462 adolescents (age 13, Infinium MethylationEPICv1 Beadchip) from Project Viva.

Batch effects were corrected using ComBat when applicable (24). Blood cell type proportions were estimated in each dataset using the reference-based Houseman method implemented in *ENmix* (22). The FlowSorted.Blood.450K and FlowSorted.Blood.EPIC references were used for each dataset according to the DNAm array employed (36,37).

Following the same procedures as in the discovery phase, we tested the association between blood cell DNAm (M-values) and asthma with exacerbations using *limma* linear robust regression models adjusted for age, sex, ancestry, and cell-type heterogeneity in each dataset separately. Ancestry was corrected using 2 PCs of WGS in GALA II, 2 PCs of genotyping data in GEMAS, and self-reported race and ethnicity in Project Viva, according to available data. Project Viva analyses were adjusted by body mass index (BMI) z-scores (38). Genomic inflation was inspected using Q-Q plots and λ_GC_, and it was detected and corrected in GALA II 450K–PR using the BACON Bayesian method (λ_Uncorrected_: 1.81, λ_Corrected_: 1.15). We meta-analyzed the replication cohorts using METASOFT, and replication was declared for CpGs showing the same direction of effect and *p*<0.05 in the meta-analysis among replication datasets.

#### Cross-tissue evalution

Genome-wide CpGs with evidence of replication were followed for cross-tissue evaluation in 393 nasal samples from Project Viva collected in early adolescence. Sample collection, DNAm measurement, and QC have been previously described (38). Briefly, nasal swabs were collected from the anterior nares during the adolescent visit (age 13). DNAm was measured using the Illumina Infinium MethylationEPICv1 Beadchip. Batch effects for sample plates were corrected using ComBat (24), adjusting for age, sex, asthma, exacerbations, and self-reported race and ethnicity. Cell-type heterogeneity was captured using the free-reference ReFACTor algorithm (39), due to the lack of reference datasets for this tissue. Robust linear models were adjusted for age, sex, self-reported race and ethnicity, BMI (z-scores), and 10 ReFACTor components. Validation was declared for CpGs showing the same direction effect and *p*<0.05.

#### Differentially methylated regions (DMRs)

DMRs were estimated in the three discovery datasets using three independent software: *dmrff* (40), *DMRcate* (41), and *ipDMR* (42). Regions were started at CpGs with *p*<0.05 (*dmrff*) or false discovery rate (FDR)<0.05 (*DMRcate* and *ipDMR*), and those CpGs located within 1000 bp were combined into the same region. All other parameters were set to their default values. For *dmrff*, we used the dmrff.meta() function to conduct the meta-analysis, including as input DNAm M-values and EWAS summary statistics (corrected by BACON) for each dataset. For *DMRcate* and *ipDMR*, we used as input the summary statistics of the discovery meta-EWAS. We retained DMRs that contain ≥2 CpGs and remain significant after multiple-testing correction at the region level (adjusted *p*<0.05; Bonferroni correction in *dmrff* and FDR correction in *DMRcate* and *ipDMR*). DMRs were annotated to the nearest gene using *rGREAT*. To minimize false-positive results, we prioritized DMRs detected using the three different software. Genomic regions were considered overlapping across software if they shared ≥50% of their genomic coordinates.

#### Cell-type deconvoluted EWAS

Genome-wide significant CpGs identified in whole blood (tissue-level bulk data) were analyzed for differential DNAm at cell-type resolution detection using the tensor compositional analysis (TCA) in R (25). We analyzed the discovery datasets, in which cell-type proportions were deconvoluted using BayesCCE (26). We used tca() for fitting the TCA model, which was modified to extract the estimates and standard errors to conduct a meta-analysis. We included age and sex as cell-type level covariates (C1) and genotype and technical PCs as tissue-level covariates (C2). We tested the association between asthma with exacerbations and cell-type-specific DNAm using a marginal conditional test in which a model was fitted jointly across all cell types, and the effect of each cell type was tested separately. For each cell type, we meta-analyzed the three ethnic groups using METASOFT (28). We applied an FDR<5% to account for multiple testing across cell types and genomel1lwide CpGs (5 cell types x 505 CpGs).

#### Cis-methylation quantitative trait loci (meQTL) analysis

WGS data were generated for African Americans and Hispanics/Latinos from SAGE and GALA II within the Trans-Omics for Precision Medicine (TOPMed) Consortia at New York Genome and Northwest Genomics Centers. Briefly, paired-end reads (2×150 bp) were generated on the HiSeq X^TM^ Ten platform (Illumina), achieving a minimum mean genome coverage of 30x. All technical and preprocessing details are described elsewhere (43).

We examined the genetic influence on DNAm at genome-wide significant CpGs through a cis-meQTL analysis. We analyzed 1,668 individuals with WGS and DNAm data independently of asthma and exacerbation status, including 721 African Americans, 572 Puerto Ricans, and 375 Mexican Americans. For each CpG site, all single-nucleotide polymorphisms (SNPs) located within ±250 kb (upstream and downstream) with a minor allele frequency (MAF)≥5% were analyzed. We tested associations between DNAm M-values and SNP genotypes using linear regression models in tensorQTL (44), adjusting for age, sex, asthma and exacerbations status, genetic ancestry, cell-type proportions, and technical PCs. Alleles were aligned to a common reference using R and PLINK 2.0 (45), and results were meta-analyzed across ethnic groups using METASOFT (28). Significance was declared using an FDR<5%, adjusting for all evaluated CpG-SNP comparisons (587,945 tested pairs). Independent meQTL signals were detected using the clumping method in PLINK 1.9 (500 kb, *r^2^*<0.2) (45), accounting for linkage disequilibrium patterns across the three ethnic groups. For each clump, the lead SNP is selected as the one with the lowest *p*-value.

#### Cis-expression quantitative trait methylation (eQTM) analysis

Whole-blood transcriptomes were generated in African Americans and Hispanics/Latinos from SAGE and GALA II using RNA-seq within the TOPMed Consortia. Data was normalized and preprocessed following the GTEx project v8 protocol as described elsewhere (46). Briefly, RNA was isolated from whole-blood samples collected in PAXgene blood RNA tubes, and 101 bp paired-end reads were generated in a HiSeq 4000 platform (Illumina). Count-level data were generated based on the GRCh38 human reference genome and the GENCODE 30 gene annotations. Gene counts were transformed into the trimmed mean of M values (TMM), and TMMs were normalized using an inverse normal transformation.

We examined associations between DNAm at genome-wide significant CpGs and nearby gene expression through a cis-eQTM analysis. We analyzed 1,209 individuals with RNA-seq and DNAm data independently of asthma and exacerbation status, including 512 African Americans, 362 Puerto Ricans, and 335 Mexican Americans. For each CpG site, all genes whose transcription start site (TSS) is located within ±250 kb (upstream and downstream) were considered in the analyses. We estimated surrogate variables to account for batch effects and unknown sources of variation in the gene expression matrix, using the *sva* R package (24). Null models were adjusted for age, sex, genetic ancestry, CBC, and technical PCs, while full models also included asthma and exacerbation status (non-asthmatics, non-exacerbator asthmatics, and exacerbators). We filtered out surrogate variables that were moderately to highly correlated (*r^2^*>0.5, *p*<0.05) with DNAm-derived cell counts to minimize collinearity in the eQTM models (**Figure S3**).

We tested associations between normalized gene expression and DNAm M-values using linear regression models in MatrixQTL (47), adjusting for age, sex, asthma and exacerbation status, genetic ancestry, cell-type proportions, technical PCs, and surrogate variables of gene expression. Analyses were conducted for each ethnic group from the discovery phase and meta-analyzed using METASOFT (28). Significance was declared using an FDR<5%, adjusting for all evaluated CpG-gene comparisons (3,766 tested pairs).

#### Enrichment analyses and druggable targets

We conducted enrichment analyses to gain potential mechanistic insights into identified epigenetic markers associated with asthma and exacerbations. First, we examined whether genome-wide CpGs associated with asthma with exacerbations were shared with other traits based on previous EWAS using the EWAS Toolkit (32).

Second, gene-set enrichment analyses (GSEA) were conducted using Enrichr to identify enrichment in biological pathways, gene ontologies, human diseases, and/or drug signatures (48). We analyzed gene annotations for suggestive associations in the discovery meta-EWAS using an arbitrary cutoff of *p*<5×10^-7^, and we queried the databases BioPlanet 2019, KEGG 2019 Human, Panther 2016, GO Biological Process 2025, DisGeNET, and DSigDB. Significant enrichment terms were declared using an FDR<0.05. To ensure the robustness of associations, we prioritized enrichment terms that remained significant (*p*<0.05) after reassessing GSEA by varying alternative input cutoffs for gene selection (*p*<5×10^-6^ and *p*<5×10^-8^).

Third, we searched for candidate druggable genes and potential therapies for asthma among approved or in-development drugs using the Drug Gene Interaction (DGldb) database (49). As input data, we used gene annotations for genome-wide significant CpGs and prioritized those drugs with the highest number of gene interactions.

#### DNAm predictors of plasma proteins and aging

The methscore() *ENmix* function was used to calculate multiple DNAm predictors, including 109 DNAm-predicted plasma protein levels and first-, second-, and third-generation epigenetic clocks (50). We analyzed clocks predictive of chronological age (HorvathAge, HannumAge, SkinBloodAge), mortality/morbidity (PhenoAge), telomere length (DNAmTL), and pace of aging (PACE). Additionally, we used the *methylCIPHER* R package to calculate the SystemsAge clock, which estimates aging in 11 physiological systems based on whole-blood DNAm (Heart, Lung, Kidney, Liver, Brain, Immune, Inflammatory, Blood, Musculoskeletal, Hormone, and Metabolic) (51).

In the discovery cohorts, we tested associations between DNAm predictors and asthma with exacerbations using linear regression models adjusted by age, sex, genetic ancestry, blood cell-type proportions, and technical PCs. Analyses were conducted separately for each discovery ethnic group and meta-analyzed employing a random-effects model (restricted maximum likelihood) with the *metafor* package in R (52). Significance was declared using an FDR<0.05, considering the number of plasma proteins or epigenetic clocks analyzed.

## RESULTS

### Study population

We analyzed 1,192 participants in the discovery phase, including 502 African Americans, 466 Puerto Ricans, and 224 Mexican Americans. Their characteristics are summarized in **Table 1** and **Table S1**. Briefly, the mean age was 13.1-15.3 years, 52% were female, and 23% were obese. We analyzed 690 controls (participants without asthma) and 502 cases (asthma patients reporting exacerbations in the past year). Among asthma subjects, 74.5% had moderate-to-severe asthma and 85% uncontrolled asthma. Mean predicted forced expiratory volume in 1 second (FEV_1_) ranged from 101.8-116.7%, predicted forced vital capacity (FVC) from 105.6-119.8%, predicted FEV_1_/FVC from 96.1-99.5%, and total serum IgE from 270-502 IU/ml.

**Table 1.**
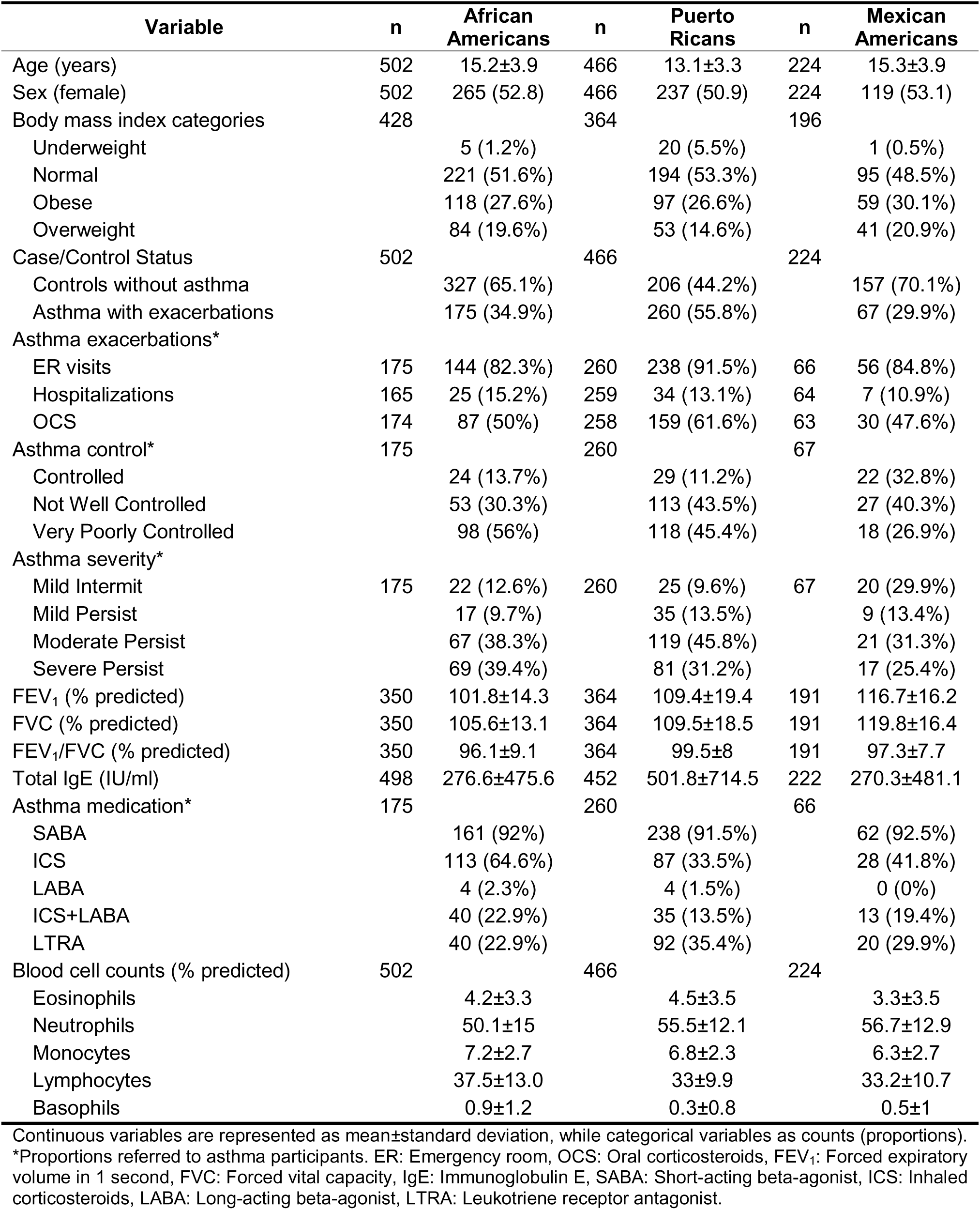
Descriptives summary of participants analyzed in the discovery phase.

| Variable | n | African Americans | n | Puerto Ricans | n | Mexican Americans |
| --- | --- | --- | --- | --- | --- | --- |
| Age (years) | 502 | 15.2±3.9 | 466 | 13.1±3.3 | 224 | 15.3±3.9 |
| Sex (female) | 502 | 265 (52.8) | 466 | 237 (50.9) | 224 | 119 (53.1) |
| Body mass index categories | 428 |  | 364 |  | 196 |  |
| Underweight |  | 5 (1.2%) |  | 20 (5.5%) |  | 1 (0.5%) |
| Normal |  | 221 (51.6%) |  | 194 (53.3%) |  | 95 (48.5%) |
| Obese |  | 118 (27.6%) |  | 97 (26.6%) |  | 59 (30.1%) |
| Overweight |  | 84 (19.6%) |  | 53 (14.6%) |  | 41 (20.9%) |
| Case/Control Status | 502 |  | 466 |  | 224 |  |
| Controls without asthma |  | 327 (65.1%) |  | 206 (44.2%) |  | 157 (70.1%) |
| Asthma with exacerbations |  | 175 (34.9%) |  | 260 (55.8%) |  | 67 (29.9%) |
| Asthma exacerbations* |  |  |  |  |  |  |
| ER visits | 175 | 144 (82.3%) | 260 | 238 (91.5%) | 66 | 56 (84.8%) |
| Hospitalizations | 165 | 25 (15.2%) | 259 | 34 (13.1%) | 64 | 7 (10.9%) |
| OCS | 174 | 87 (50%) | 258 | 159 (61.6%) | 63 | 30 (47.6%) |
| Asthma control* | 175 |  | 260 |  | 67 |  |
| Controlled |  | 24 (13.7%) |  | 29 (11.2%) |  | 22 (32.8%) |
| Not Well Controlled |  | 53 (30.3%) |  | 113 (43.5%) |  | 27 (40.3%) |
| Very Poorly Controlled |  | 98 (56%) |  | 118 (45.4%) |  | 18 (26.9%) |
| Asthma severity* |  |  |  |  |  |  |
| Mild Intermittent | 175 | 22 (12.6%) | 260 | 25 (9.6%) | 67 | 20 (29.9%) |
| Mild Persistent |  | 17 (9.7%) |  | 35 (13.5%) |  | 9 (13.4%) |
| Moderate Persistent |  | 67 (38.3%) |  | 119 (45.8%) |  | 21 (31.3%) |
| Severe Persistent |  | 69 (39.4%) |  | 81 (31.2%) |  | 17 (25.4%) |
| FEV<sub>1</sub> (% predicted) | 350 | 101.8±14.3 | 364 | 109.4±19.4 | 191 | 116.7±16.2 |
| FVC (% predicted) | 350 | 105.6±13.1 | 364 | 109.5±18.5 | 191 | 119.8±16.4 |
| FEV<sub>1</sub>/FVC (% predicted) | 350 | 96.1±9.1 | 364 | 99.5±8 | 191 | 97.3±7.7 |
| Total IgE (IU/ml) | 498 | 276.6±475.6 | 452 | 501.8±714.5 | 222 | 270.3±481.1 |
| Asthma medication* | 175 |  | 260 |  | 66 |  |
| SABA |  | 161 (92%) |  | 238 (91.5%) |  | 62 (92.5%) |
| ICS |  | 113 (64.6%) |  | 87 (33.5%) |  | 28 (41.8%) |
| LABA |  | 4 (2.3%) |  | 4 (1.5%) |  | 0 (0%) |
| ICS+LABA |  | 40 (22.9%) |  | 35 (13.5%) |  | 13 (19.4%) |
| LTRA |  | 40 (22.9%) |  | 92 (35.4%) |  | 20 (29.9%) |
| Blood cell counts (% predicted) | 502 |  | 466 |  | 224 |  |
| Eosinophils |  | 4.2±3.3 |  | 4.5±3.5 |  | 3.3±3.5 |
| Neutrophils |  | 50.1±15 |  | 55.5±12.1 |  | 56.7±12.9 |
| Monocytes |  | 7.2±2.7 |  | 6.8±2.3 |  | 6.3±2.7 |
| Lymphocytes |  | 37.5±13.0 |  | 33±9.9 |  | 33.2±10.7 |
| Basophils |  | 0.9±1.2 |  | 0.3±0.8 |  | 0.5±1 |

Continuous variables are represented as mean±standard deviation, while categorical variables as counts (proportions).
\*Proportions referred to asthma participants. ER: Emergency room, OCS: Oral corticosteroids, FEV<sub>1</sub>: Forced expiratory volume in 1 second, FVC: Forced vital capacity, IgE: Immunoglobulin E, SABA: Short-acting beta-agonist, ICS: Inhaled corticosteroids, LABA: Long-acting beta-agonist, LTRA: Leukotriene receptor antagonist.

### Epigenome-wide association study of asthma with severe exacerbations

In the multi-ethnic meta-EWAS, we identified 505 CpGs (90% hypomethylated) genome-wide associated with asthma with severe exacerbations (*p*<9×10^-8^, λ_Corrected_: 1.05, **Figure 2A**, **Table 2**, **Figure S4**, **Table S3**). Compared to previous EWAS of asthma-related traits in any tissue, we reported novel associations for 423 CpGs and 190 genes (**Figure S5**). The top-hit hypomethylated CpG was annotated to *HTT* (cg04078283, logFC: -0.24, *p*=4.7×10^-15^) and the top-hit hypermethylated to *SLAMF1* (cg18881723, logFC: 0.37, *p=*5.9×10^-14^). We identified genes implicated in cytokine signaling (*IL18RAP*, *IL23R*, *JAK1*, *TGFB1*, *TGFBR3*, *TNFAIP8L1*, *TNFSF4, CXCL12*), innate immunity (*DICER1, PRF1, GZMB*, *GZMH*), asthma treatment response (*ADRB2*, *TPST2*), and insulin pathway (*IGF1R*, *IGF2R*), and genes located in the 17q12–q21 locus (*CCL18*, *ACACA*, *CNP*, *MAPT*, *LRRC37A*, *TBX21*, *LRRC46*, *ABI3*, *PHB1*, *FAM117A*) and paralogs of genes associated with asthma exacerbations (*APOBEC3H*, *CACNA2D2*, *ZBTB18*).

**Figure 2.**
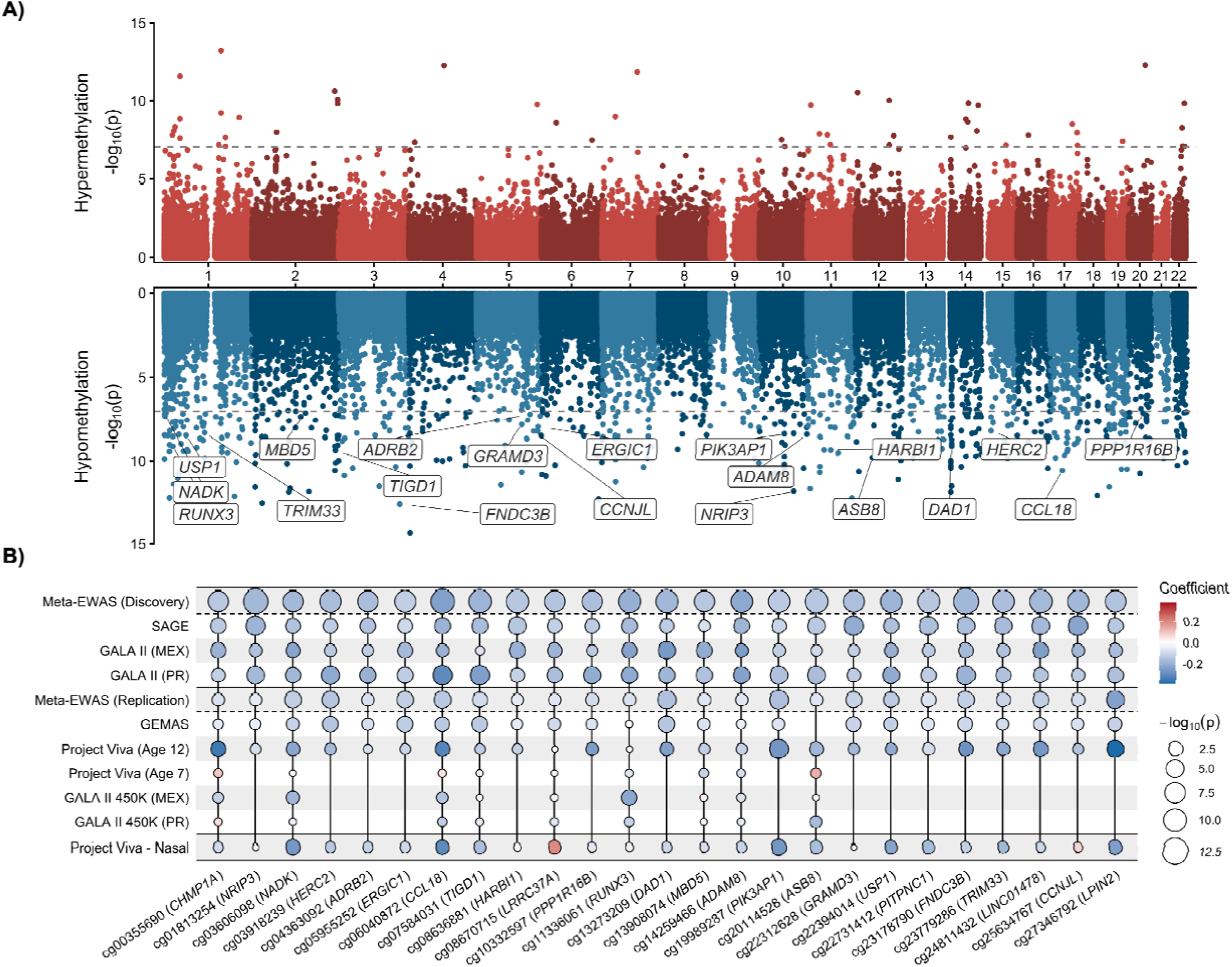
**A)** Miami plot of the epigenome-wide meta-analysis of asthma with exacerbations in the discovery phase. Association -log_10_(*p*) is represented on the y-axis and genomic coordinates on the x-axis. CpGs hypermethylated in cases are represented in the upper panel (red) and those hypermethylated in the lower panel (blue). The gray dashed line represents the genome-wide significance threshold (*p*<9x10^-8^). Gene annotation is provided for CpGs with evidence of replication. **B)** Bubble plot showing the summary statistics across the discovery, replication, and cross-tissue evaluation stages for those CpGs with evidence of replication. Positive associations are shown in red and negative associations in blue. Bubble size is proportional to -log_10_(*p*). Missing bubbles indicate CpGs not available in one or more datasets due to quality control filtering or array coverage.

**Table 2.**
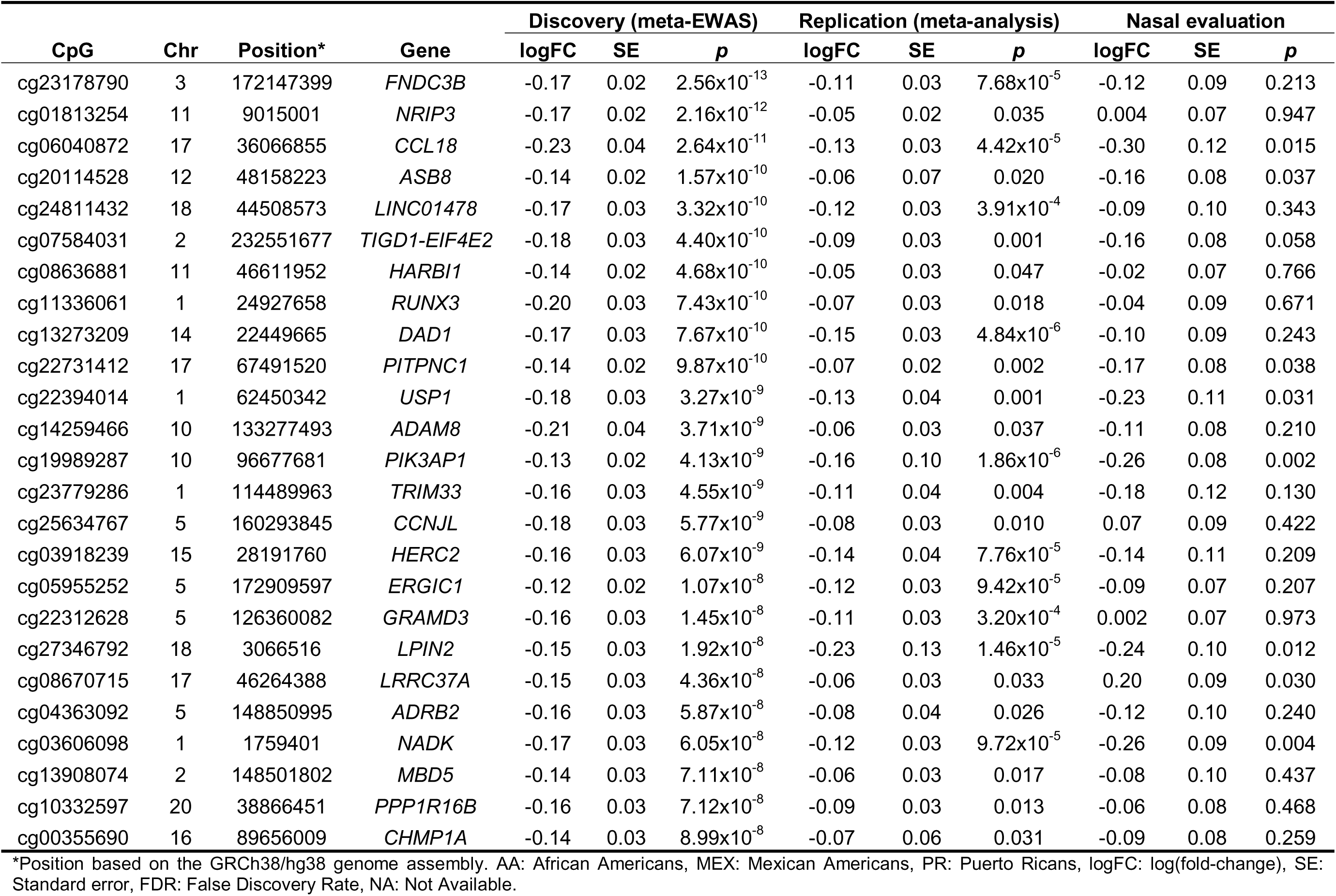
Summary table of genome-wide CpG sites with evidence of replication.

### Replication analyses

Genome-wide CpGs were followed for replication in a meta-analysis among 1,516 participants from diverse ethnicities and age groups, including 979 Europeans/White, 326 Hispanics/Latinos, 133 Black, and 78 other or mixed ethnic groups (**Table S1**). Among the 505 CpG sites tested, 25 CpGs replicated in the meta-analysis with the same effect direction (*p*<0.05, **Figure 2B**, **Table 2, Figure S6**, **Table S4**), including genes in the 17q12-q21 locus (*CCL18*, *LRRC37A*) and involved in treatment response (*ADRB2*), immunity (*RUNX3*, *PIK3AP1*, *LPIN2*, *TRIM33*), airway inflammation and remodeling (*ADAM8*), and ubiquitin-specific processing (*USP1*, *ASB8*).

### Cross-tissue evaluation in nasal epithelia

The 25 CpG replicated CpGs in blood cells were examined in 393 nasal samples from ethnically diverse adolescents (**Table S1**). A total of 8 CpGs were associated with asthma and exacerbations in nasal epithelium, of which 7 showed the same direction effect as in whole blood (*p*<0.05, **Figure 2B**, **Table 2**), including CpGs annotated to *PIK3AP1*, *NADK*, *LPIN2*, *CCL18* (17q12), *USP1*, *ASB8*, and *PITPNC1*.

### Regional methylation analyses

We identified 119 DMRs (83% hypomethylated) associated with asthma with exacerbations in the discovery cohorts using three independent software (FDR<0.05, **Figure 3A-B**, **Tables S5-S8**). These included genes involved in T-cell activity (*BACH2*, *BCL11B*, *RUNX3*, *ID2*, *CLEC16A, TBX21*), chemokine and cytokine signaling (*CCL4*, *CX3CR1, IL18RAP*, *TNFRSF1A*, *TNFRSF4*, *NMUR1*), airway inflammation (*PTPN6*, *PIK3R2*), and NF-kB signaling (*CARD11*), and some located in the 17q12-q21 locus (*CCL4*, *TBX21*, *CNP*, *STH*). The top-hit hypomethylated DMR was annotated to *CLEC16A* (logFC: -0.23, *p*=4.6×10^-17^) and the top-hit hypermethylated to *CHD3* (logFC: 0.17, *p*=8.6×10^-23^). Some DMRs were annotated to the same genes as replicated CpGs (*DAD1*, *LPIN2*, *PITPNC1*, *RUNX3*).

**Figure 3.**
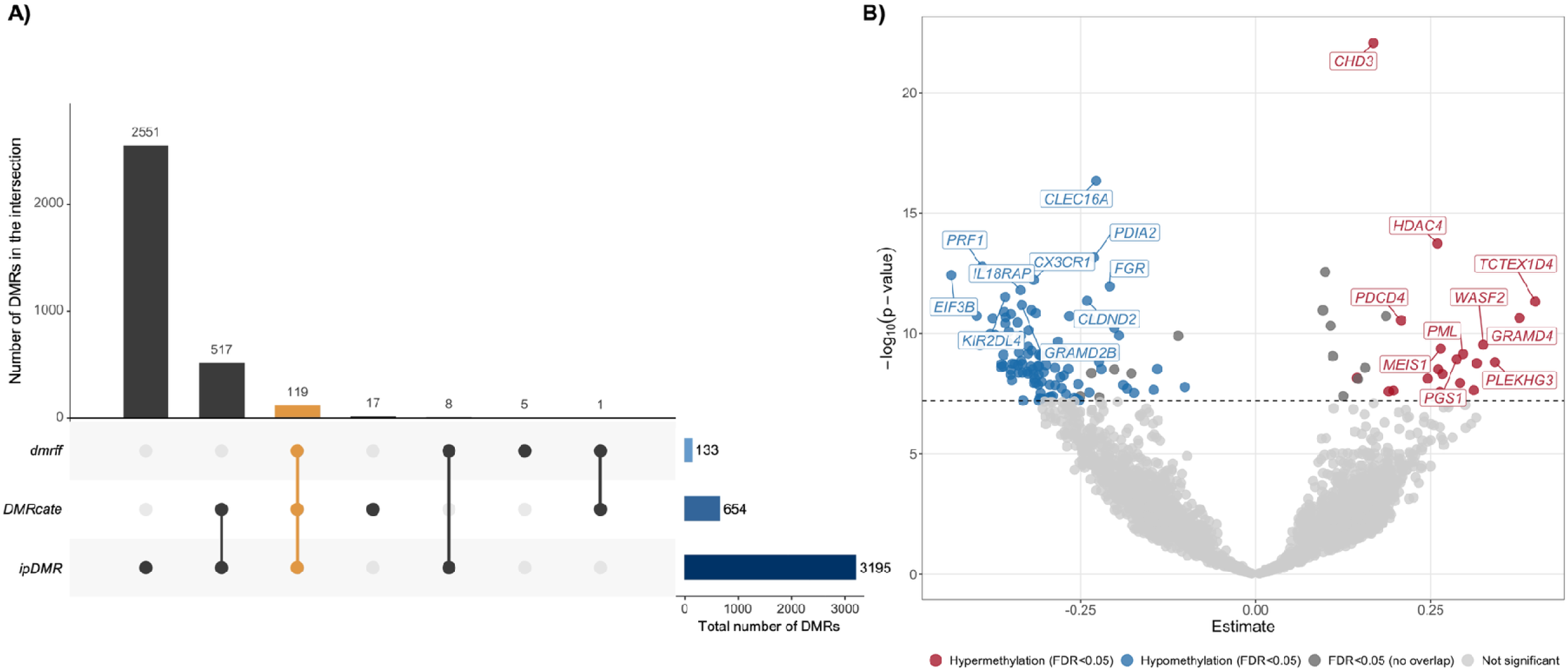
**A)** UpSet plot showing the overlap among differentially methylated regions (DMRs) detected by three independent methods (*dmrff*, *DMRcate*, and *ipDMR*). The horizontal bars on the left show the total number of DMRs detected by each software. The vertical bars indicate the number of DMRs in each intersection (the intersection among the three software is highlighted in orange). **B)** Volcano plot of detected DMRs associated with asthma with exacerbations using *dmrff*. Association -log_10_(*p*) is represented on the y-axis and effects on the x-axis. Only those significant DMRs (FDR<0.05) detected with the three different software are colored in blue (hypomethylation) and in red (hypermethylation). Gene annotation is provided for the top ten most significant hypo- and hyper-methylated DMRs.

### Leukocyte cell-type deconvoluted EWAS

Across the 505 significant CpGs at whole-blood bulk DNAm, we identified 42 CpGs with cell-type-specific DNAm effects (FDR<0.05, **Figure 4A**, **Table S9**). Most of the signals were identified in lymphocytes (n=32; e.g., *FNIP2*, *HTT*, *PIK3AP1*, *CD300A*, *PLCG2*), the second most abundant cell type (**Table 1**, **Figure 4B**), in addition to neutrophils (n=8; e.g., *CCL18*, *PIK3AP1*, *APOBEC3H*), monocytes (n=2; *CLDND2*, *ADAP1*), and/or basophils (n=2; *PLEKHG3*, *CNR2*). We observed differences in cell-type-specific signatures for the genome-wide CpGs across ethnic groups (**Figure 4C**). At the nominal significance threshold, lymphocytes were the cell type contributing the most to observed associations in African Americans (39%), whereas neutrophils predominated in Puerto Ricans (47%). No cell-type-specific signature was observed in Mexican Americans.

**Figure 4.**
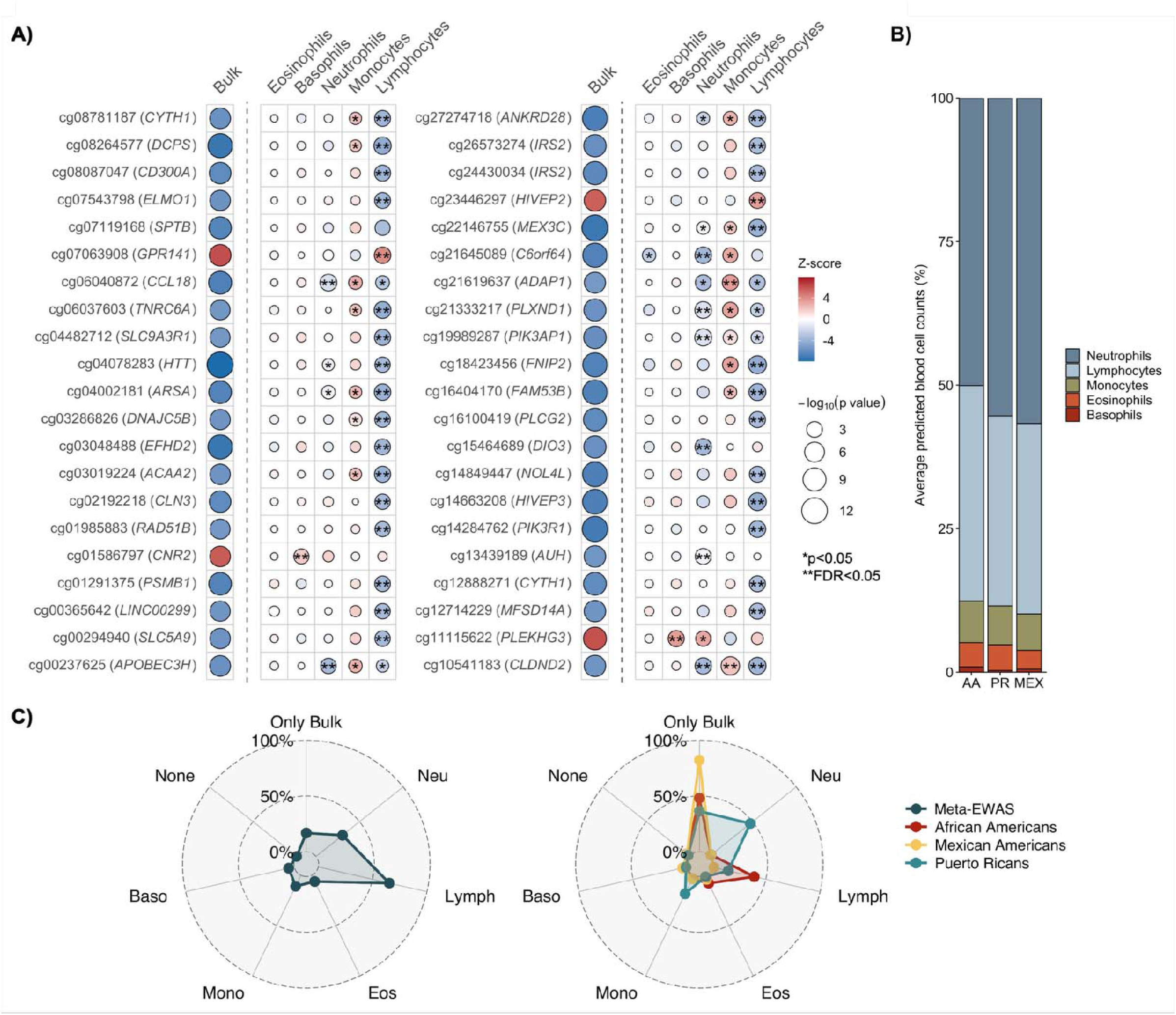
**A)** Bubble plot showing the summary statistics in the cell-type deconvoluted meta-analyses for those CpGs showing any significant cell-type-specific DNAm association. Positive associations are shown in red and negative associations in blue. Bubble size is proportional to -log_10_(*p*). Coefficients were normalized to Z-scores (estimate/SE) and reported for the whole-blood bulk DNAm analysis and for the conditional marginal test for each blood cell type. \**p*<0.05. **FDR<0.05. **B)** Bar plot of the average predicted blood cell type proportions in each ethnic group (AA: African Americans, PR: Puerto Ricans, MEX: Mexican Americans). **C)** Radar chart showing the proportion of CpGs showing cell-type-specific nominal associations (*p*<0.05) among the genome-wide CpGs identified in the discovery meta-EWAS. The meta-analysis is shown in the left plot, and the ethnic-stratified analyses are shown in the right plot.

### Integration with gene expression and genetic variation through quantitative trait loci analyses

We identified 134 CpGs acting as cis-eQTM in whole blood, analyzing 1,209 participants from the discovery (290 CpG-Gene significant pairs, FDR<0.05, **Figure 5A**, **Table S10**), including 118 eQTM associations with T-cell receptor genes, in addition to genes involved in NK/T cell signaling. We found associations with several genes involved in inflammation, cytokine and chemokine signaling, and/or asthma (*IFNAR1*, *TBX21*, *TNFRSF4, TNFRSF14*, *TNFRSF18*, *NMUR1,* and *CX3CR1*). At seven of the 25 replicated CpGs, blood DNAm levels were associated with whole blood gene expression of *SKAP1*, *HDAC7*, *GFOD1*, *DENND2B*, *TRAV19*, *NFE2L1*, *NMUR1*, and *AP4B1*.

**Figure 5.**
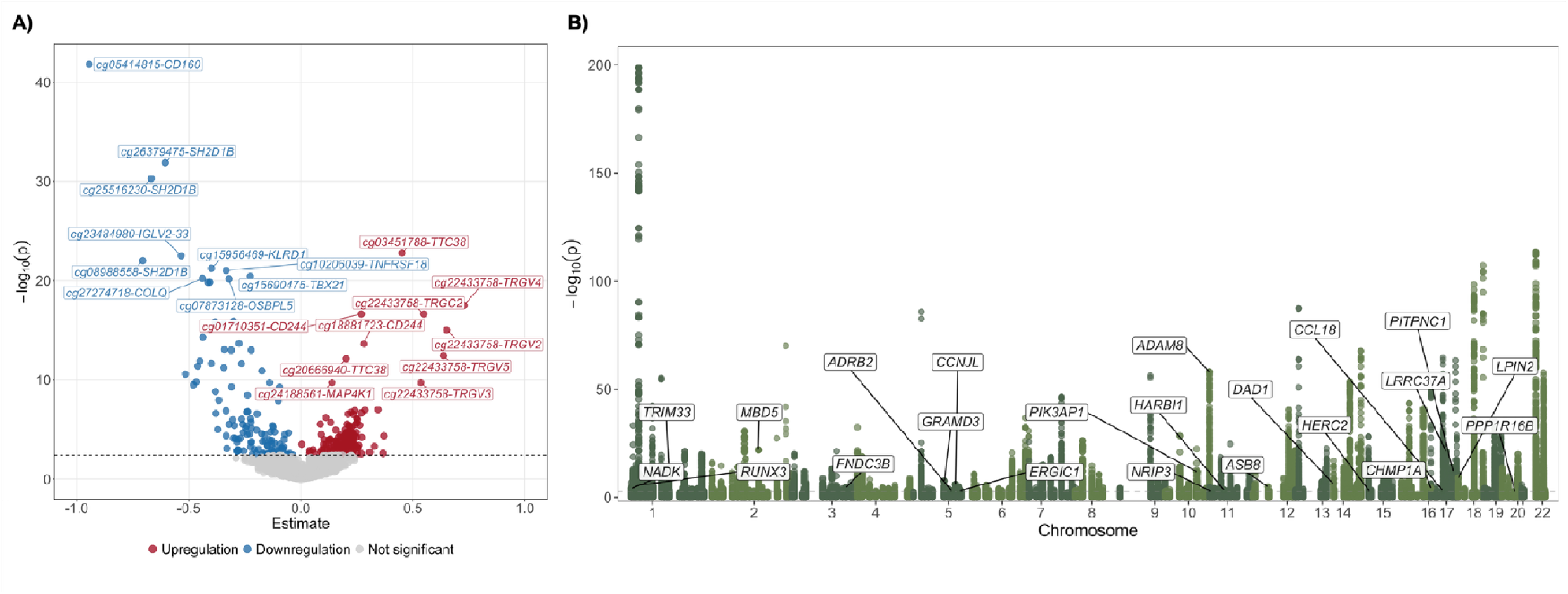
**A)** Volcano plot of CpG-gene pairs analyzed in the cis-expression quantitative trait methylation (eQTM) meta-analysis. Association -log_10_(*p*) is represented on the y-axis and effects on the x-axis. Only those significant eQTMs (FDR<0.05) are colored in blue (negative association) and in red (positive association). Gene annotation is provided for the top ten most significant CpGs associated with down- and upregulation of nearby genes. **B)** Manhattan plot of the association of genetic variants with genome-wide CpGs through a methylation quantitative trait loci (meQTL) meta-analysis. Each dot corresponds to a single genetic variant tested as a meQTL. Association -log_10_(*p*) is represented on the y-axis and genomic coordinates on the x-axis. The gray dashed line represents the FDR<0.05 significance threshold. Gene annotation is provided for the top-hit meQTL for CpGs with evidence of replication.

Moreover, 88% of the genome-wide CpGs (446 out of 505 CpGs) were associated with at least ≥1 independent meQTL in 1,668 blood samples of discovery cohorts (28,137 significant SNP-CpG pairs, 2,673 independent meQTLs, FDR<0.05, **Figure 5B**, **Table S11**). Top-hit association was for the SNP rs6424130 acting as meQTL of cg01586767 (*CNR2*, β_C allele_: 0.82, *p*=2.0×10^-199^). Additionally, CpG cg18455083 (*MBP*) had the highest number of independent meQTLs (n=44) associated with its DNAm levels. CpGs with evidence of replication were associated with genetic variation, except 3 CpGs annotated to *TIGD1*-*EIF4E2*, *USP1*, and *LINC01478*. Among the 25 replicated CpG sites, the cg14259466 (*ADAM8*) reported the greatest number and most significant meQTL associations (17 independent meQTLs; top-hit: rs2995312, β_T allele_: 0.40, *p*=1.0×10^-58^).

### Enrichment analyses

CpGs genome-wide associated with asthma and exacerbations in the discovery phase were associated by previous EWAS with environmental exposures (air pollution exposure and smoking) and asthma-related traits (fractional exhaled nitric oxide, atopy, and childhood/allergic asthma) (**Figure 6A**, **Table S12**). GSEA showed an overrepresentation of genes among discovery CpGs implicated in IL-2, T and B cell activation, Th1/Th2/Th17 cell differentiation, insulin/insulin-like growth factors (IGF), and vascular endothelial growth factor (VEGF) signaling pathways (**Figure 6B**, **Table S13**). We detected an enrichment of genes associated with immune disorders (e.g., arthritis) and likely regulated by drugs in development (e.g., trichostatin A).

**Figure 6.**
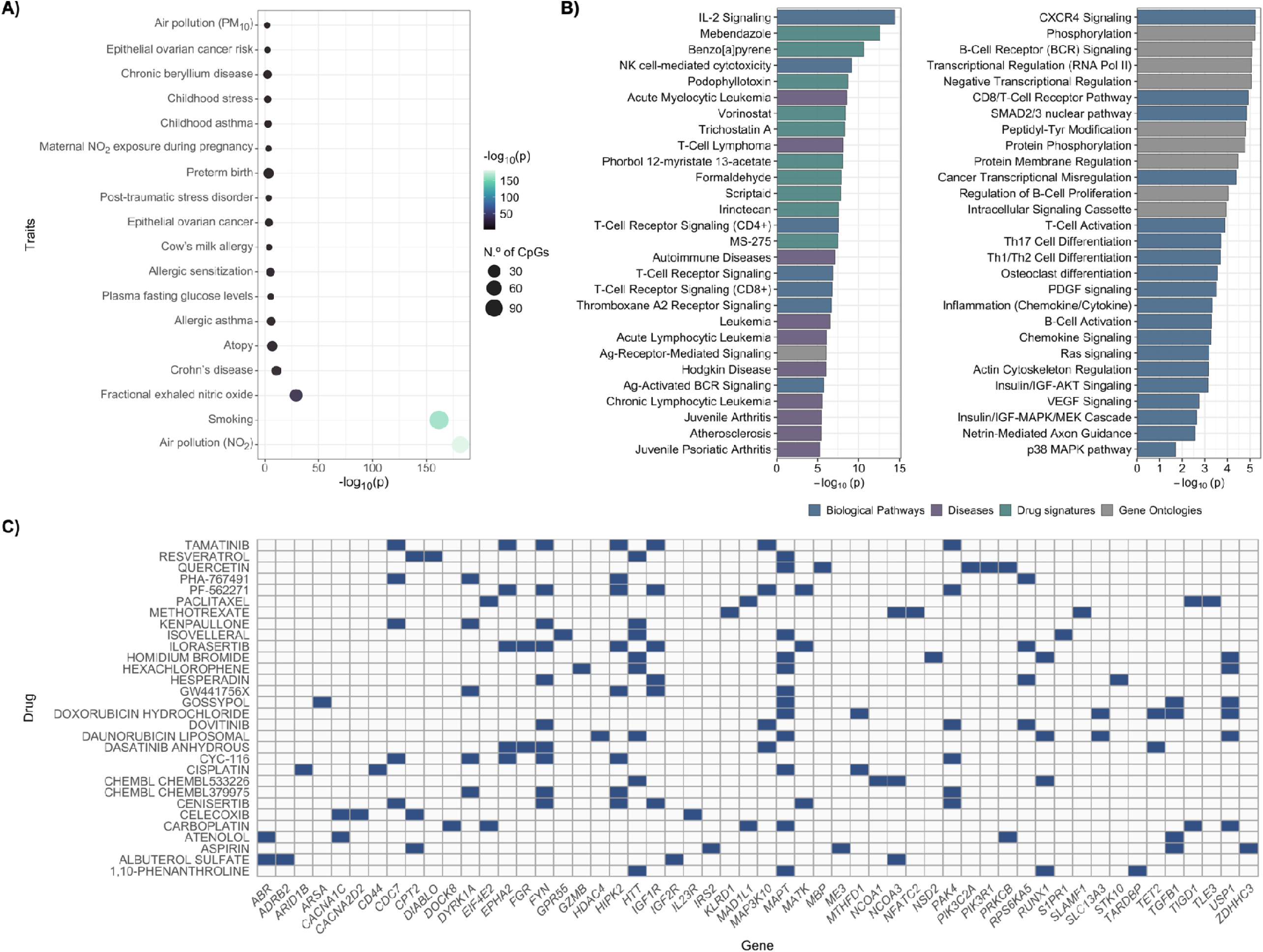
**A)** Bubble plot summarizing the CpG-based enrichment analyses in other traits by previous EWAS using EWAS toolkit. The enrichment terms are represented on the y-axis and the -log_10_(*p*) of the enrichment on the x-axis. Bubble size is proportional to the number of CpGs detected for each trait and colored by -log_10_(*p*). **B)** Bar plots summarizing the gene-set enrichment analysis using Enrichr. The enrichment terms are represented on the y-axis and the -log_10_(*p*) of the enrichment on the x-axis. Bars are colored according to the category of the term. Only the top ten most significant terms of each queried database are represented (FDR<0.05). Terms identifiers were simplified for graphical representation. If any term overlaps between different databases, only the most significant one is plotted. **C)** Heatmap showing potential drug-gene interactions detected using DGldb. Only the top 30 drugs showing the highest number of gene interactions are represented.

### Search for druggable genes

Among the 505 genome-wide CpGs from the discovery, 120 were potential druggable genes by approved and/or in-development drugs (2,268 drug-gene interactions) (**Table S14**). These included several bronchodilators, anti-inflammatories, and antiallergic drugs already approved for asthma and/or related diseases. Tamatinib and ilorasertib were the two developmental drugs with the highest number of potential gene interactions, in addition to other drugs like resveratrol, quercetin, albuterol, and aspirin (**Figure 6C**, **Table S14**).

### Associations with plasma protein level predictors

DNAm-predicted plasma levels of 21 proteins were associated with asthma with exacerbations across admixed and minoritized children (FDR<0.05, **Figure 7A**, **Table S15**). These included proteins involved in inflammation (IL-19, SERPINA3, CCL22, Complement C5a, and chemerin), airway remodeling and/or transforming growth factor (TGF-β) pathway (metalloproteinase 1 [MMP-1], myostatin, and BMP1), host defense (Granzyme A, Granulysin, Complement C9, CD169, lactoferrin, PIGR, and CCL25), and glucose metabolism (insulin receptor, chemerin, and FGF21).

**Figure 7.**
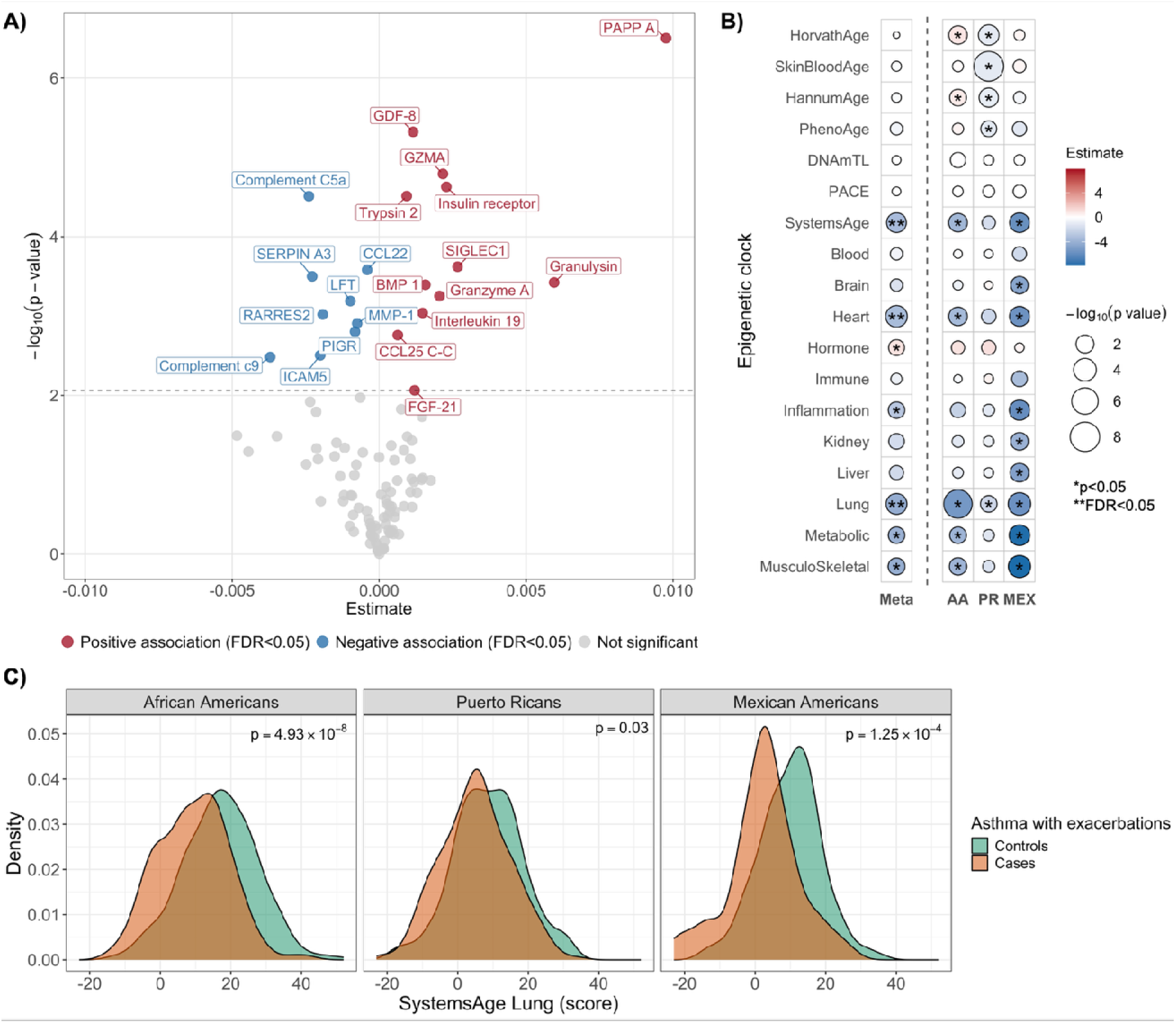
**A)** Volcano plot of DNAm-predicted plasma proteins associated with asthma and exacerbations. Association -log_10_(*p*) is represented on the y-axis and effects on the x-axis. Only those significant proteins (FDR<0.05) are colored in blue (negative association) and in red (positive association). **B)** Bubble plot showing the summary statistics of the association of epigenetic clocks with asthma and exacerbations. Positive associations are shown in red and negative associations in blue. Bubble size is proportional to -log_10_(*p*). Statistics are represented for the meta-analysis and ethnic-stratified analyses (AA: African Americans, PR: Puerto Ricans, MEX: Mexica Americans). \**p*<0.05. **FDR<0.05. **C)** Density plots of the SystemsAge Lung score for participants without asthma (green) and for those with asthma and exacerbations (orange). P-values indicate the association between the clock and asthma with exacerbations, stratified by ethnic group.

These proteins showed a significant protein-protein interaction network (*p*=4.4x10^-6^) and grouped into four clusters (**Figure S7**). These protein networks were enriched for humoral and antimicrobial responses, interactions with viral proteins, and inflammatory and immune responses (FDR<0.05, **Table S16**).

### Epigenetic aging

All clocks except PACE showed moderate-to-high correlations with chronological age among discovery ethnic groups (**Figures S8-S10**). The SystemsAge clock was significantly associated with asthma with exacerbations (FDR<0.05, **Figure 7B**, **Table S17**). We detected negative associations for aging scores for the heart, lung, inflammation, metabolic, and musculoskeletal physiological systems, and a positive association for the hormone clock. The lung clock (SystemsAge) was consistently negatively associated across the three ethnic groups (*p*-values range between *p*≥4.9×10^-8^ and *p*≤0.03, **Figure 7C**). Classical epigenetic clocks did not show consistent and significant effects across ethnic groups (**Figure 7B**, **Table S17**).

## DISCUSSION

This is the first multi-ancestry meta-EWAS of extreme asthma phenotypes, in which we detected 505 CpGs and 119 DMRs in minoritized and admixed populations. We replicated the association between extreme asthma phenotypes and blood cell DNAm at 25 CpGs in ethnically diverse cohorts, of which 7 were cross-tissue validated 7 in nasal epithelium. Cell-type deconvolution and eQTM analyses supported that T-cells primarily drive the epigenetic signature of asthma. Identified CpGs are likely to be susceptible to genetic variation due to meQTLs and environmental exposures to air pollution and smoking. We identified 120 potentially druggable genes, suggesting novel candidates for asthma therapies. Finally, we associated asthma and exacerbations with 21 DNAm-predictors of plasma proteins involved in antimicrobial, inflammatory, and immune responses, and a lung epigenetic aging clock.

Consistent with prior whole-blood meta-EWAS (19,53,54), most asthma-associated CpGs were hypomethylated. We identified loci with strong genetic evidence of association with asthma (e.g., the 17q12-21 locus and genes such as *ADRB2*, *TBX21*, or *ADAM8*), supporting the robustness of our analyses. In addition to 16,692 CpGs associated with asthma or exacerbations to date (2,150 CpGs in whole blood), our multi-ancestry meta-EWAS of asthma with exacerbations identified 423 CpGs previously undetected in any tissue (32–35). The extreme-comparison design and larger sample size enabled the detection of hundreds of novel signals compared to a previous EWAS of asthma in the same discovery populations (19). Gene annotations indicate that most of our DNAm signals involve immune and inflammatory processes and airway pathophysiology.

We observed enrichment of genes in the ILl1l2 signaling pathway, genomel1lwide CpGs and DMRs annotated to IL-18 and IL-23 receptors (*IL18RAP*, *IL23R*), and higher DNAm-predicted plasma levels of IL-19 in subjects with asthma and exacerbations. These pro-inflammatory ILs, dysregulated in asthma, are involved in NK cell activation, antibody responses, and T-cell biology. IL-2 is central to T-cell proliferation and differentiation, IL-18 to Th1/Th2 balance and interferon-γ (IFN-γ) responses, and IL-23 to Th17 response (55). Targeting IL pathways (IL-4, IL-5) is the mechanism of action of current biological therapies that are successful in treating asthma (56). Although the anti-IL-23 antibody risankizumab has been investigated in severe asthma, further studies are needed to assess its efficacy for specific phenotypes (56–58). The importance of ILs among detected DNAm markers was supported by a replicated CpG acting as eQTM of neuromedin U receptor 1 (*NMUR1*), which plays a key role in IL production and T2 inflammation in asthma (59).

Furthermore, we identified several DNAm markers annotated to genes or associated with their expression that are implicated in the tumor necrosis factor-alpha (TNF-α) pathway (*TNFAIP8L1*, *TNFRSF1A*, *TNFSF4*, *TNFRSF14*, and *TNFRSF18*). *TNFRSF4* (OX40/CD134) participates in the anti-inflammatory effects of OCS and is the target of the monoclonal antibody amlitelimab, which is under evaluation in phase IIa trials for severe asthma (58,60,61). Although modulation of the TNFl1lα pathway remains of interest, clinical trials of TNF inhibitors (e.g., etanercept, infliximab) have shown limited efficacy in asthma (58). Some genes identified in our meta-EWAS (*IL23R*, *KLRD1*, and *NFATC2*) are potential targets of anti-TNF drugs, suggesting that these loci could play a role in the relationship between these drugs and asthma. Other identified loci (*TNFRSF14* and *TNFAIP8L1*) and family-related proteins have been implicated in airway inflammation and hyperresponsiveness and represent additional candidate targets (62,63).

Moreover, we detected DNAm markers related to TGF-β signaling (*TGFB1* and *TGFBR3*) and DNAm-predicted increases in TGF-β members/activators (myostatin and BMP-1) in asthma patients. TGF-β modulates T-cell differentiation, suppressing Treg cells and promoting Th17 responses, and is a central mediator of airway remodeling (55,64). We also identified DNAm markers linked to MMPs, which contribute to airway remodeling and represent potential therapeutic targets (58,65). We replicated one CpG on *ADAM8*, a disintegrin-and-metalloprotease implicated in severe asthma whose inhibition reduces airway hyperresponsiveness and Th2 inflammation in mice (66). Although MMP-1 is often increased in the airways of severe asthma patients (58,65), we observed lower DNAm-predicted serum MMP-1 in exacerbators, which may relate to steroid therapies (61).

Additionally, we identified multiple epigenetic loci (*CXCL12*, *CCL18*, *CCL4*, and *CX3CR1*) and DNAm-predicted proteins (CCL22 and CCL25) implicating chemokine signaling. A CpG annotated to *CCL18* was replicated, validated in nasal tissue, and showed neutrophill1lspecific differential DNAm. *CCL18* is highly expressed in lungs and has been associated with lung function by genetic studies (67). Elevated CCL18 plasma levels throughout childhood have also been described as a risk factor for asthma (68). In the 17q12-q21 locus, the genetic signal most robustly associated with childhood asthma (3), we found CpGs associated with asthma with exacerbations acting as eQTM of *TBX21*, *NFE2L1*, and *SKAP1*. *TBX21* encodes a transcription factor involved in IFN-γ signaling and airway hyperresponsiveness, suggested as a candidate pharmacogenetic gene (69). *NFE2L1* is a family member of the nuclear factor erythroid 2-related factor 2 (*NFE2L2*), whose inhibition reduces airway inflammation and neutrophil infiltration (58). *SKAP1* participates in T cell activation and represents a potential therapeutic target for autoimmune and inflammatory diseases (70).

Notably, using a hypothesis-free approach, we detected a CpG annotated to *ADRB2* (69,71). Although this gene has been extensively studied in relation to asthma treatment response, its efficacy as a pharmacogenetic biomarker is uncertain (69,71). Our findings suggest that, in addition to SNPs, *ADRB2* DNAm markers might be of interest for asthma precision medicine. Additionally, we detected genes, or their paralogs, previously associated with treatment response and/or asthma exacerbations by genomic and epigenomic studies (*TPST2*, *APOBEC3H*, *CACNA2D2*, and *ZBTB18*) (34,35,72–74).

Our results suggest that T cells primarily underlie the DNAm signature of asthma and exacerbations, in agreement with previous evidence for asthma and allergies (75). Celll1ltype deconvolution showed that most signals were lymphocytel1ldriven, supported by 112 eQTM associations with Tl1lcell receptor genes and gene-enrichment of Th1/Th2/Th17 pathways. Statistical power of cell-type deconvolution methods is expected to correlate with cell-type proportions (25). The fact that strong signals in TCA arise in T cells (i.e., not the most abundant cells), along with supporting RNA-seq eQTM evidence, suggests that these signals are particularly robust. Furthermore, we observed ancestryl1lspecific differences in cell-type contributors, highlighting lymphocytes in African Americans and neutrophils in Puerto Ricans. Indeed, different racial/ethnic variations in airway inflammation have been detected based on clinical variables (76).

Furthermore, we observed enrichment of insulin/IGF pathways, including CpGs annotated to *IGF1R* and *IGF2R*, and DNAm-predicted increases in insulin receptor and metabolic regulators (chemerin and FGF21). Obesity and insulin resistance are risk factors for poor asthma control and exacerbations (77). Our results align with prior evidence supporting potential repurposing of anti-diabetic agents for asthma exacerbation management (14,77), whose benefits may relate to DNAm modifications. In addition, we observed enrichment for genes likely regulated by trichostatin A, a potential asthma drug highlighted by genomic studies (71,72).

Enrichment analyses indicated that identified CpGs may reflect environmental exposures to smoking and air pollution, which are risk factors for asthma and exacerbations, partially through DNAm changes (9). However, meQTL analyses also supported a genetic influence, with 88% of the CpGs associated with genetic variation. Notably, CpGs with the strongest meQTL signals did not replicate, suggesting that population-specific genetic architectures may influence DNAm effects (20,78). These findings support that gene-environment factors contribute to the asthma DNAm signature.

Moreover, we identified 120 druggable genes, with tamatinib and ilorasertib showing the highest number of potential gene interactions. Tamatinib (R406) is a developmental drug for asthma that prevents IgE-mediated and allergen-induced airway hyperresponsiveness, airway inflammation, and mast cell degranulation (79–81). Ilorasertib, primarily evaluated in cancer, is a VEGF pathway inhibitor, which may justify further investigation given the VEGF-pathway upregulation in asthma (82). Additionally, we highlighted quercetin and resveratrol, antioxidant flavonoids of therapeutic interest for respiratory diseases owing to their antiviral and anti-inflammatory properties, which can be comparable to those of budesonide (83). Although these are exploratory analyses that warrant further research, they are supported by the identification of asthma-related drugs (e.g., albuterol, formoterol, mometasone, and prednisone).

Finally, DNAm-predictions suggested dysregulation of plasma proteins involved in inflammatory and antimicrobial responses (e.g., granzymes), implicating NK cell and innate immune activation genes. These DNAm modifications could impact microbial infection susceptibility, the major triggers of asthma exacerbations (3,84). Additionally, the unexpected lung-aging deceleration (SystemsAge) in youth with asthma could reflect an immaturity of lung development (85). However, this association may be biased by clinical (e.g., corticosteroid effects on DNAm) (7), technical (e.g., heterogeneity between our populations and those where SystemsAge was trained, which comprised mostly non-Hispanic White adults) (51), or other unmeasured factors. Although other chronological DNAm clocks have shown unexpected associations with allergic traits (85,86), potentially reflecting greater health-seeking behavior among children with chronic conditions (86), the relationship between this lungl1lage clock and asthma requires further validation.

Our study has several strengths. First, it represents the largest discovery meta-EWAS of asthma to date, conducted with the EPICv1 array, providing 57-68% greater genomic coverage of analyzed CpGs than prior meta EWAS using the 450K array (53,54). Second, we addressed genomic diversity by analyzing four ethnic/ancestral groups, including diverse and historically underrepresented populations. Third, we applied state-of-the-art statistical methods to ensure the robustness of our findings, including correction for known/unknown confounders and genomic inflation, multiple comparison adjustment, and providing replication and cross-tissue evaluations. Fourth, we conducted extensive multi omic characterization, including cell type deconvolution, WGS and RNA seq integration, pathways and drugs identification, and DNAm based protein and aging predictors.

Nevertheless, some limitations should be acknowledged. First, cross-sectional analyses do not identify molecular mechanisms and causality. Although DNAm markers were assumed to exert cis-regulatory effects, their regulatory effects may be more complex. Second, replication may be limited by a lack of independent cohorts with similar ancestry and EPICv1 data, as well as asthma exacerbation data, reducing our ability to replicate population-specific markers. Third, our cell-type-resolution analyses relied on *in-silico* methods, and single-cell assays are required for in-depth characterizations. Fourth, microarrays only covered ∼3% of CpGs across the epigenome (87), with other epigenetic markers remaining unexamined (75).

## CONCLUSIONS

The first meta-EWAS of extreme asthma phenotypes identified hundreds of novel blood DNAm markers. Several loci involved in bronchodilation, immunity, and airway inflammation and remodeling were replicated and cross-tissue validated in nasal samples. Blood cell-type deconvolution and integration with RNA-seq supported a key role for T cells in asthma DNAm signature. Our study suggested candidate drugs for asthma treatment and dysregulation of DNAm-predicted plasma proteins involved in host defense and a lung epigenetic clock in asthma patients.

## Data Availability

All summary statistics of all analyses supporting the main findings and conclusions of this manuscript are reported in the main text, supplementary material, and/or Zenodo repository (accession number 10.5281/zenodo.18637179). Summary statistics of genome-wide CpGs identified in the discovery meta-analysis will be openly published at the EWAS Atlas after manuscript acceptance. TOPMed whole-genome sequencing data and RNA-seq data from GALA II and SAGE are available in the database of Genotypes and Phenotypes (dbGaP) under accession numbers phs000920.v4.p2 and phs000921.v4.p1, respectively. Due to ethical restrictions and the consent provided by participants, demographic and epigenetic data from the analyzed cohorts are not publicly available. Access may be granted on reasonable request directed to the corresponding and/or senior authors of this article, and subject to approval by the principal investigators of the studies and ethics committees.

## AUTHOR CONTRIBUTIONS

JP-G, AC, and MP-Y were involved in the conceptualization of the study; JP-G, ER, AC, and MP-Y in methodology; JP-G, EM-G, ZJC, and AG in formal analysis and investigation; JW, FL-D, RG-P, JMH-P, PP-G, EM-L, IS-M, JR-S, JV, MF-H, EO, DRG, LNB, EZ, EGB, AC, and MPY in resources; JP-G, EM-G, MM-A, CE, AB, JRE, DH, and SH in data curation; JP-G in visualization; JP-G, AC, and MP-Y in data interpretation and writing the original draft; EGB, AC, and MP-Y in funding acquisition; and AC and MP-Y in project supervision. All authors were involved in the critical revision and editing of the manuscript and have read and agreed to the published version. The authors agree to be accountable for all aspects of the work, ensuring its scientific accuracy and integrity.

## DATA SHARING STATEMENT

All summary statistics of all analyses supporting the main findings and conclusions of this manuscript are reported in the main text, supplementary material, and/or Zenodo repository (accession number 10.5281/zenodo.18637179). The datasets on the Zenodo repository will be openly available after manuscript acceptance and available for the reviewers through the peer review process using the following <u>link</u>. Summary statistics of genome-wide CpGs identified in the discovery meta-analysis will be openly published at the EWAS Atlas after manuscript acceptance. TOPMed whole-genome sequencing data and RNA-seq data from GALA II and SAGE are available in the database of Genotypes and Phenotypes (dbGaP) under accession numbers phs000920.v4.p2 and phs000921.v4.p1, respectively. Due to ethical restrictions and the consent provided by participants, demographic and epigenetic data from the analyzed cohorts are not publicly available. Access may be granted on reasonable request directed to the corresponding and/or senior authors of this article, and subject to approval by the principal investigators of the studies and ethics committees.

## ACKNOWLEDGMENTS

We acknowledge the patients, families, recruiters, healthcare providers, and/or community clinics for their participation in all the analyzed cohorts (GALA II, SAGE, GEMAS, and Project Viva). We thank Sandra Salazar for her support as GALA II study coordinator. We thank the investigators and teams at the New York Genome Center and the Northwest Genomics Center for WGS sample preparation, quality control, and data generation and processing. We gratefully acknowledge the studies and participants who provided biological samples and data, and those researchers who have contributed to the NHLBI Trans-Omics for Precision Medicine (TOPMed) Consortium (https://www.nhlbiwgs.org/topmed-banner-authorship).

## ABBREVIATIONS

BMI: Body mass index
CpG: 5′-cytosine-phosphate-guanine-3′ dinucleotides
DNAm: DNA methylation
eQTM: Expression quantitative trait methylation
EWAS: Epigenome-wide association studies
FDR: False discovery rate
FEV_1_: Forced expiratory volume in 1 second
FVC: Forced vital capacity
GDF-8: Growth differentiation factor 8 or myostatin
GSEA: Gene-set enrichment analyses
IFN-γ: Interferon-γ
IgE: Immunoglobulin E
IGF: Insulin-like growth factors
IL: Interleukin
LFT: Lactotransferrin or lactoferrin
meQTL: Methylation quantitative trait loci
MMP: Metalloproteinase
PC: Principal component
RARRES2: Retinoic acid receptor responder protein 2 or chemerin
SNP: Single nucleotide polymorphism
TGF-β: Transforming growth factor
TNF-α: Tumor necrosis factor-alpha
VEGF: Vascular endothelial growth factor
WGS: Whole-genome sequencing

