## Supplementary Material for "Multi-Ancestry Epigenome-Wide Meta-Analysis Identifies Novel Bulk and Cell-Type-Specific Epigenetic Markers of Asthma with Severe Exacerbations"

**SUPPLEMENTARY METHODS**

**Study Populations**

*Genes-environments & Admixture in Latino Americans study (GALA II)*: Case-control study of pediatric asthma among Latino children from the United States (US) (1). Briefly, children and young adults (aged 8-21 years old) who self-identified as Hispanic/Latino and had four Hispanic/Latino grandparents were recruited between 2006 and 2014 through clinical and community-based recruitment centers in different regions of the USA and Puerto Rico. Asthma cases were defined by 1) physician diagnosis of asthma, 2) recent use of asthma controller/reliever medication, or 3) occurrence of ≥2 asthma symptoms (cough, wheeze, or shortness of breath) in the two years before enrollment. Individuals were excluded according to the following exclusion criteria: 1) history of other lung or chronic illnesses other than atopy and allergy-related diseases, 2) any smoking within one year of recruitment date, 3) ≥10 pack-years of smoking, and/ or 4) women in the third trimester of pregnancy. GALA II was approved by the Human Research Protection Program (HRPP) Institutional Review Board (IRB) at the University of California, San Francisco (UCSF) (UCSF-IRB No. 10-00889). In this study, we analyzed whole-blood samples from 466 Puerto Ricans and 224 Mexican Americans in the discovery phase, and 84 independent Puerto Ricans and 159 Mexican Americans in the replication phase.

*Study of African Americans, Asthma, Genes & Environments (SAGE)*: Case-control study of pediatric asthma conducted in parallel with GALA II, except that participants identified as African American and reported having four African American grandparents (1). SAGE was approved by the HRPP-IRB of UCSF (UCSF-IRB No. 10-02877). In this study, we analyzed whole-blood samples from 502 African Americans in the discovery phase.

*Project Viva*: Prospective pre-birth cohort study of mothers and their offspring initially recruited between 1999 and 2002 during the first prenatal visits at Atrius Harvard Vanguard Medical Associates, a multispecialty medical group practice in Massachusetts (US) (2,3). Project Viva was launched in 1999 to study the influences of environmental and social exposures during the perinatal period on short- and long-term health outcomes in mothers and their offspring, mainly including pregnancy outcomes, maternal mental and cardiometabolic health, and child neurodevelopment, asthma/atopy, and obesity/cardiometabolic health. Eligibility criteria included fluency in English, gestational age <22 weeks at the first prenatal visit, planned to deliver in one of the designated study hospitals, and a singleton pregnancy. Mothers and their offspring were followed up in in-person interviews during the pregnancy until the present. The Institutional Review Board of Harvard Pilgrim Health Care reviewed and approved all study protocols. In this study, we analyzed 358 and 462 whole blood samples collected at age 7 and 13, respectively, in the replication phase. We also analyzed 393 nasal swabs collected at age 13 in the cross-tissue evaluation stage.

*Genomics and Metagenomics of Asthma Severity (GEMAS):* Ongoing case-control study aimed at investigating the molecular basis of asthma exacerbations (4). Briefly, asthma patients were recruited between 2018 and 2024 in several Allergy and Pulmonology hospital departments in Spain. Inclusion criteria for asthma participants were: 1) men and women aged 8-85 years, 2) asthma diagnosis according to the Global Initiative for Asthma (GINA) guideline, and 3) receiving asthma treatment according to GINA therapeutic steps 1-5. Patients were excluded if they had 1) ≥1 grandparents of non-European origin, 2) pregnancy, 3) coexistence of other chronic respiratory diseases, or 4) known first- or second-degree familial relationships with other participants included in the study. A group of controls aged ≥18 years old without asthma and similar demographic characteristics were recruited in a hospital allergy department after being attended for suspected drug allergies and getting a negative result. GEMAS has been approved by the ethics committees of the participating centers (approval 29/17 for the hospitals in the Canary Islands). In this study, we analyzed 453 whole blood samples from adults recruited in the Canary Islands in the replication phase.

**Clinical assessment in the discovery populations**

Demographic and clinical characteristics related to asthma development were recorded in individuals from GALA II and SAGE through standardized questionnaires. Body mass index (BMI) was categorized based on z-scores (<19 years) or BMI cutoffs (≥19 years) into underweight, normal weight, overweight, or obese based on the World Health Organization (WHO) guidelines (5).

Spirometry was conducted using a KoKo® PFT Spirometer (nSpire Health Inc., Louisville, CO), according to American Thoracic Society (ATS) recommendations as previously described (6). Predicted percentages of lung function measurements were calculated according to the Global Lung Initiative (GLI) 2012 reference equations using the *rspiro* R package (7).

Asthma control and severity were defined according to the 2007 National Asthma Education and Prevention Program (NAEPP) Guidelines (8). We used the Childhood Asthma Control Test (C-ACT) and the Asthma Control Questionnaire (ACQ), along with medication use and lung function, to derive asthma control and severity variables. Asthma control was assessed across six domains: daytime symptoms, nighttime symptoms, activity limitations, short-acting β-agonist (SABA) use during the prior two weeks, lung function, and oral systemic corticosteroid (OCS) prescription within the past week. The five domains were categorized into three levels: controlled, not well controlled, and very poorly controlled. Asthma control was classified as the maximum category across domains, corresponding to the least favorable domain.

Asthma severity was derived from a symptom-based score and an additional medication regimen domain. The symptom-based severity score comprised five domains: nighttime symptoms, daytime symptoms, activity limitations, use of rescue medication during the past two weeks, and lung function. Each domain was categorized as mild persistent, moderate persistent, or severe persistent. The medication-based severity domain was classified into four levels according to: 1) use of SABA; 2) monotherapy with an inhaled corticosteroid (ICS), leukotriene receptor antagonist (LTRA), or theophylline; 3) combination therapy with more than one of ICS, LTRA, or theophylline; or 4) OCS prescription within the past week. Asthma severity was categorized as mild intermittent, mild persistent, moderate persistent, or severe persistent based on the maximum of the medication-based severity (four levels) and the symptom-based severity (three levels). When medication-based or symptom-based severity was discordant with asthma control, the asthma control classification was permitted to up-classify severity by one category.

Total plasma IgE was measured in duplicate using the Uni-Cap technology (Pharmacia, Kalamazoo, Mich), as previously described (9).

**Main outcome**

The main outcome of this study was asthma with severe exacerbations. Controls were defined as those participants without a medical record of asthma, asthma symptoms, use of asthma medications, or any other chronic respiratory condition. Cases were defined as those participants with asthma who had reported severe exacerbations in the past year.

Asthma was defined in GALA II and SAGE by 1) physician diagnosis of asthma, 2) recent use of asthma controller/reliever medication, or 3) occurrence of ≥2 asthma symptoms (cough, wheeze, or shortness of breath) in the two years before enrollment. In Project Viva, current asthma was defined as the mother’s report of 1) a physician diagnosis of asthma since birth, and 2) wheeze or asthma medication in the past year before enrollment. Participants reporting asthma during infancy (age 7) but without an asthma diagnosis at the early-teen visit were excluded from this study. In GEMAS, asthma was defined by a physician diagnosis based on GINA and the use of asthma medication.

In all the studies, severe asthma exacerbations were defined based on the presence of emergency care, hospitalizations, and/or OCS prescription in the past year due to asthma (10). Participants with asthma but without reporting severe exacerbations in the past year were not included in the main EWAS.

**Genome-wide DNAm measurement and quality control in the discovery phase**

In GALA II and SAGE, genomic DNA was extracted from whole-blood EDTA samples using the Wizard Genomic DNA Purification Kits 76 (Promega, Fitchburg, WI). DNAm was measured in 856,154 CpGs across the genome using the Infinium Illumina MethylationEPICv1 BeadChip (Illumina, San Diego, CA) following the manufacturer’s protocols. Samples were measured in two main batches. A standardized quality control (QC) of DNAm data was conducted using *ENmix* and *ewastools* R packages (11,12).

Briefly, bad-quality DNAm data points were defined by detection *p*>1×10^-6^ or detection beads <3. We filtered out low-quality CpGs and samples. Low-quality CpGs were those with ≥5% of bad-quality data points across samples. Low-quality samples were defined as having 1) ≥5% of bad-quality data points across probes, 2) total bisulfite intensity <3 standard deviations than bisulfite controls, and/or 3) outliers of bisulfite intensity or beta-value distribution. We corrected the background noise using the out-of-band (oob) method, the dye bias using the Regression on Logarithm of Internal Control (RELIC) method, and the probe design type bias using the Regression on Correlated Probes (RCP) method, and inter-array intensities variation by quantile normalization.

Beta values were computed from bisulfite intensities, and outlier beta values (3*IQR rule) were set as missing data. We filtered out probes and samples with a missingness rate >5% and >10%, respectively, and imputed remaining missing values (k-nearest neighbor method).

We removed potential cross-sample contaminated samples, defined as 1) discordance between the reported sex and predicted sex based on sex chromosomes DNAm, and/or 2) mixed distribution of beta values for the 59 SNP genotyping control probes, outliers from the expected distributions for AA/AB/BB genotypes. Additionally, we filtered out these control probes and those annotated to sex chromosomes (Chr X and Y). Related and duplicated individuals detected from whole-genome sequencing data ($\hat{\text{p}}$>0.2) were excluded.

Moreover, we filtered out potentially problematic probes. These included CpGs 1) without annotation in the GRCh38/hg38 build based on Illumina’s manifest, 2) related to manufacturer issues according to Illumina’s manifest, 3) with a multimodal distribution of beta values, 4) with off-target binding sites (cross-reactive probes), and 5) potentially polymorphic. Polymorphic probes were defined as those where the DNAm signal might be biased by the presence of a common single nucleotide polymorphism (SNP) with a minor allele frequency (MAF)≥5%. These were identified specifically for the individuals from GALA II and SAGE who passed the QC of DNAm using WGS data. We filtered out probes with 1) SNP at the CpG site (both C and G nucleotides), and/or 2) SNP at a single base extension from the CpG site (Type I probes). The number of samples and CpGs that passed QC is summarized in **Supplementary Table S2**.

**Genome-wide DNAm measurement and quality control in the replication phase**

*GALA II*: Sample collection was done with participants from the discovery phase. Independent subsets of 84 Puerto Ricans and 159 Mexican Americans with genome-wide DNAm data measured using the Illumina InfiniumMethylation 450K Beadchip were included in the replication phase. QC of DNAm data was conducted similarly to the discovery datasets, as previously described (13)

*GEMAS*: Blood samples were collected in EDTA tubes, as previously described. DNAm was generated using the InfiniumMethylationEPIC v1 and v2 Beadchips. QC of DNAm data was performed separately for each array version, following a similar protocol previously described for the EPICv1 subset (14). Briefly, we filtered out low-quality samples and CpGs with a >5% of missingness rate, potential cross-sample contaminated samples, and potentially polymorphic probes. QC of EPICv2 also included the removal of 1) flagged probes and probes with mapping inaccuracies based on the Illumina MethylationEPICv2.0 A2 manifest, 2) probes capturing somatic mutations (*nv* probes), and 3) replicate probes. Following a recommended framework for filtering replicate probes, we prioritized retaining the replicate with the same probe sequence as in EPICv1, and in the case of CpGs only available in EPICv2, the superior probe or the probe with the lowest root mean square deviation (RMSD) compared to whole-genome bisulfite sequencing. Data from both array versions were merged after preprocessing, and batch effects were corrected using ComBat while adjusting for asthma, age, sex, and genetic ancestry. After the QC, we analyzed 453 samples in this study (restricting to those who meet the case-control definition).

*Project Viva*: Blood samples were collected in EDTA tubes at different time points in this longitudinal pre-birth cohort. In this study, we analyzed DNAm data measured at mid-childhood (age 7) and adolescence (age 13) as described below. In both datasets, batch effects for sample plates were corrected using ComBat, adjusting for age, sex, asthma, exacerbations, and self-reported race and ethnicity. For known kinship relationships, one sample from each related pair was excluded.

- Mid-childhood: DNAm was measured using the Infinium Methylation450K Beadchip. DNAm measurement and QC have been previously described (15). After the QC, we analyzed 358 (restricting to those who meet the case-control definition).
- Adolescence: DNAm was measured in 688 samples using the Infinium MethylationEPICv1 Beadchip. DNAm data were preprocessed using the R *minfi* package. Background noise and dye-bias were corrected using the noob, probe-type correction using the beta-mixture quantile normalization (BMIQ). Samples were excluded if they 1) had a mean detection *p*>0.001, 2) had low median intensity values, 3) had a mismatch between recorded and predicted sex, 4) had a genotype mismatch if DNAm was measured for the same child at a previous timepoint (birth, early childhood, or mid-childhood), 5) had missing ID information, and/or 6) were replicates. Probes were filtered out if they were 1) probes with detection *p*>0.05 for > 1% of samples, 2) non-CpG probes, 3) probes annotated to sex chromosomes, 4) cross-reactive, and/or 5) potentially polymorphic. Potentially polymorphic and cross-reactive probes were detected using the rmSNPandCH() function in the *DMRcate* package. Additionally, polymorphic probes were detected using the *MethylToSNP* package (reliability score>0.5) or were listed as collocated with a SNP in the manifest provided by Illumina. After the QC, 462 samples were analyzed in this study (restricting to those who meet the case-control definition).

**SUPPLEMENTARY FIGURES**

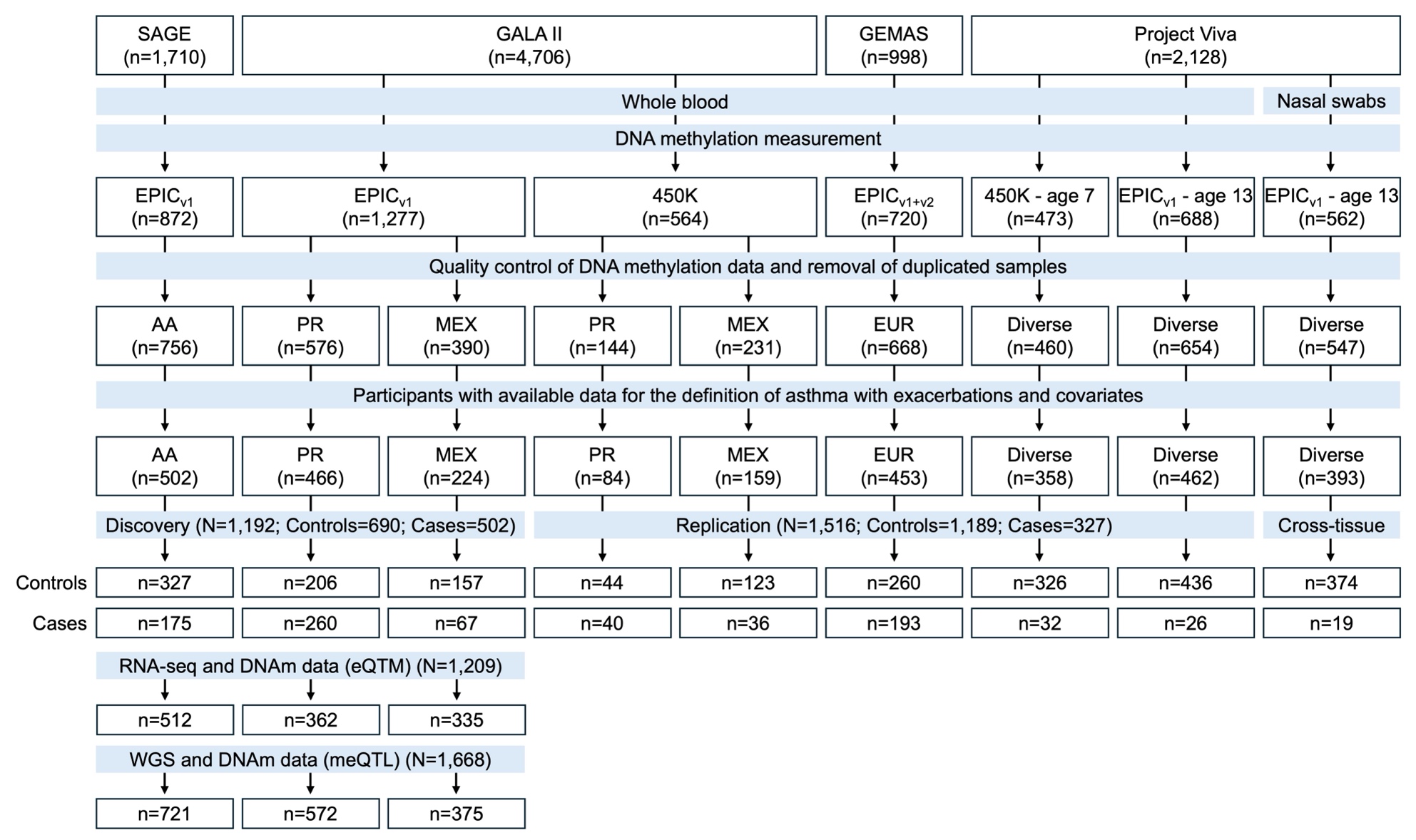

**Figure S1**. Flowchart of sample selection for the analyzed datasets.

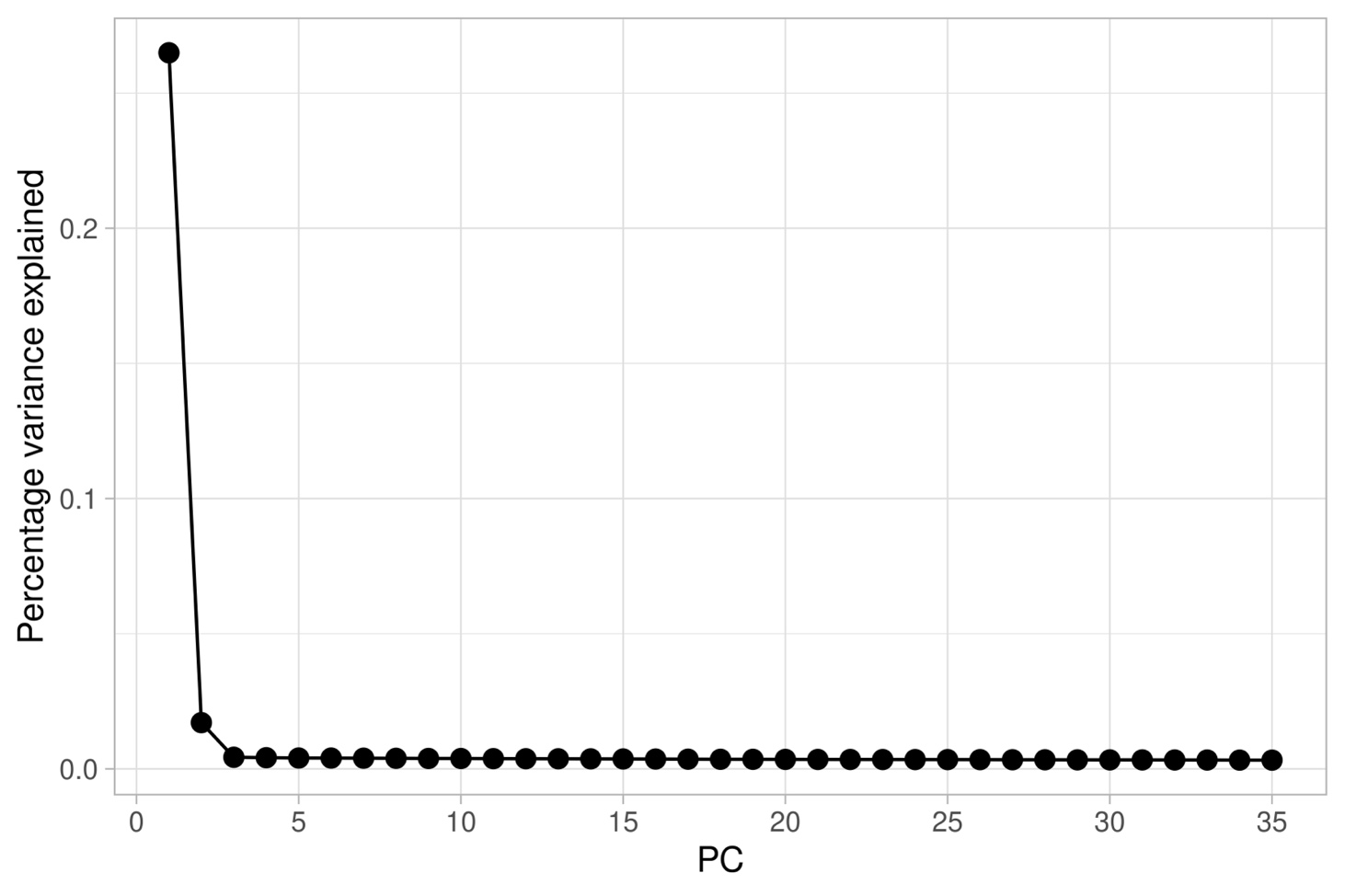

**Figure S2**. Scree plot of the percentage variance explained by the first 35 principal components of negative control probes of the Illumina Infinium MethylationEPICv1 in the discovery populations. The principal component analysis was conducted on the residuals of beta values of the negative control probes after regressing for the known technical batch.

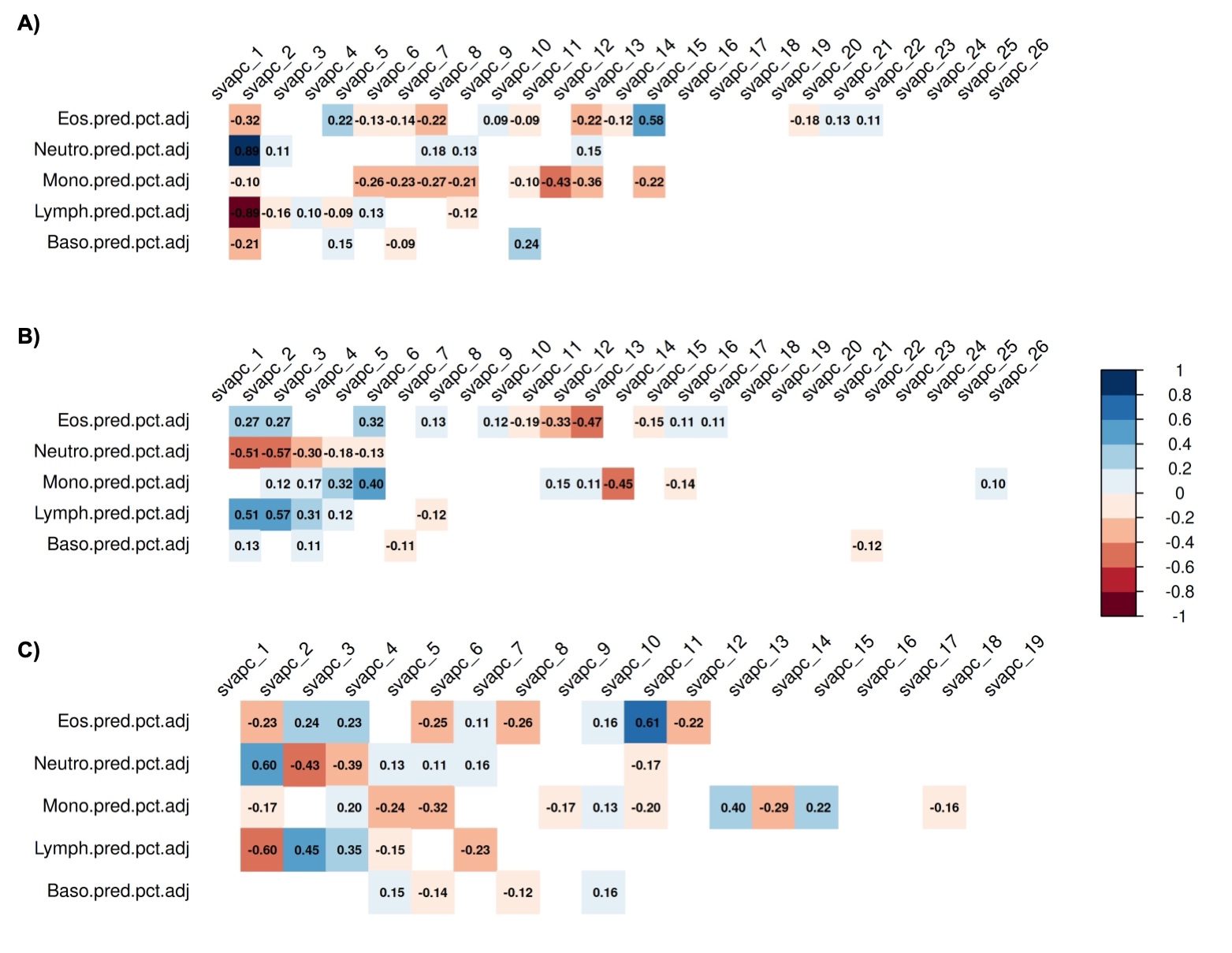

**Figure S3.** Corrplots showing the correlation among DNAm-predicted blood cell counts (BayesCCE estimations) and surrogate variables of the gene expression matrix in **A)** African Americans from SAGE, **B)** Puerto Ricans from GALA II, and **C)** Mexican Americans from GALA II. Positive correlations are shown in blue and negative associations in red. Only significant correlations are plotted (*p*<0.05). Those surrogate variables showing moderate-to-high correlations (*r^2^*<0.5, *p*<0.05) with any DNAm-predicted cell proportions were discarded as covariates in the expression quantitative trait methylation (eQTM) to minimize collinearity.

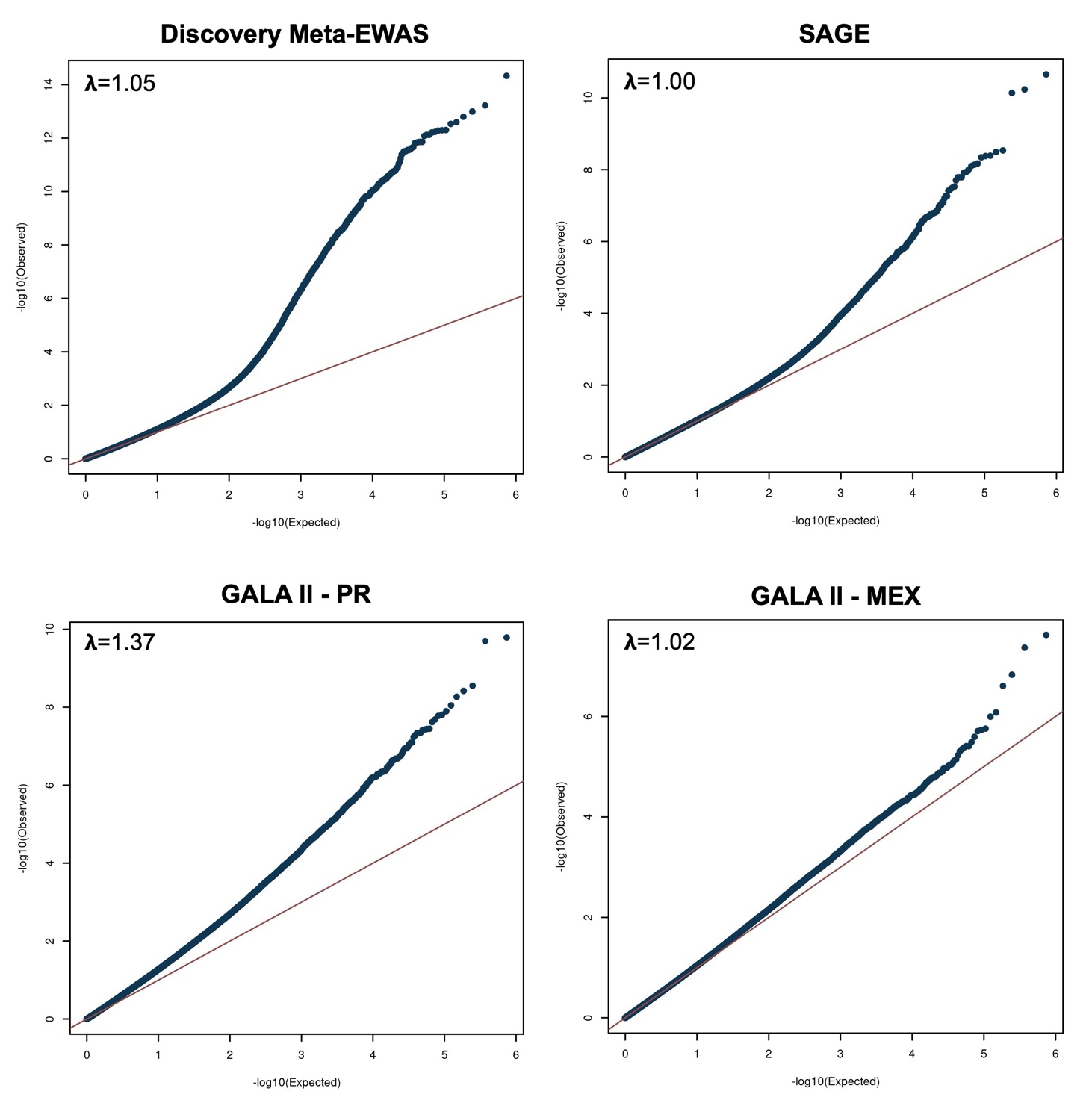

**Figure S4**. Quantile-Quantile (Q-Q) plots for epigenome-wide meta-analysis in the discovery phase and individual discovery datasets. Bias and genomic inflation were corrected for the discovery meta-analysis using the Bayesian BACON method. For individual datasets, uncorrected Q-Q plots and genomic inflation factors are shown.

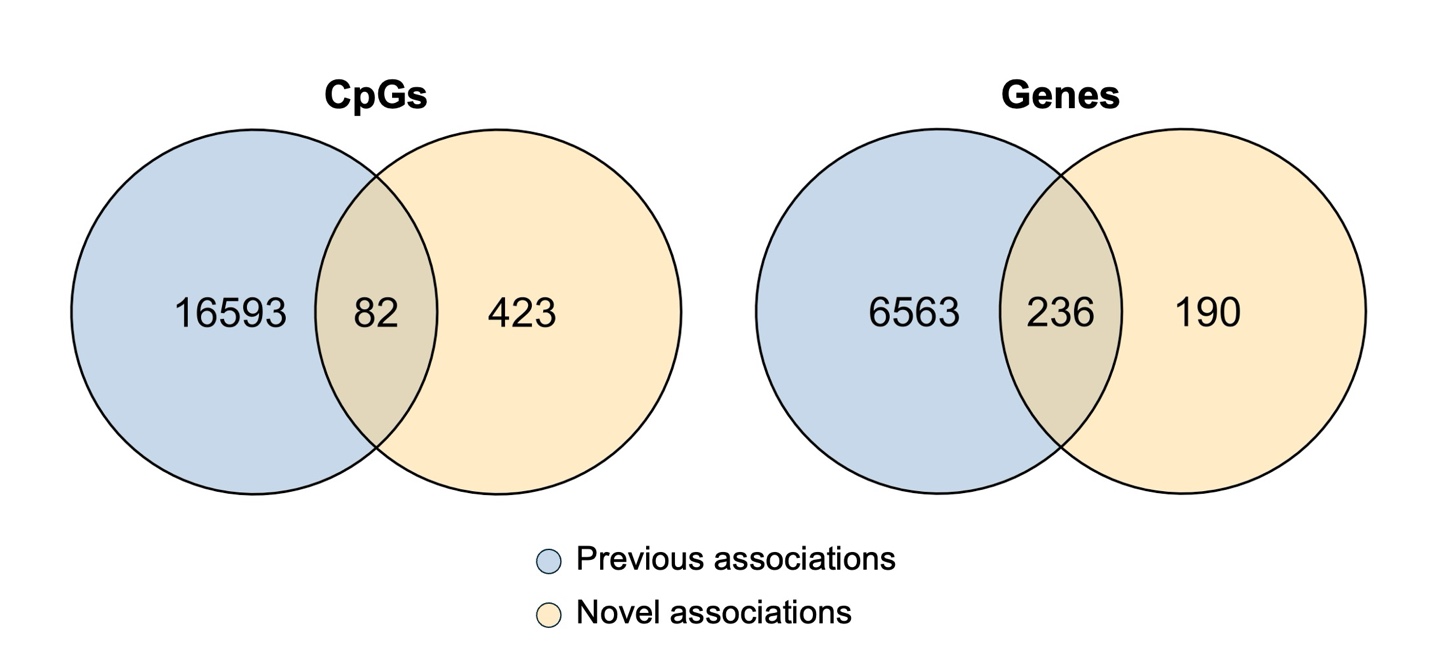

**Figure S5.** Venn diagram showing the overlap of CpGs and genes associated in our discovery meta-analysis of asthma with exacerbations and those previously reported by EWAS of asthma and/or exacerbations.

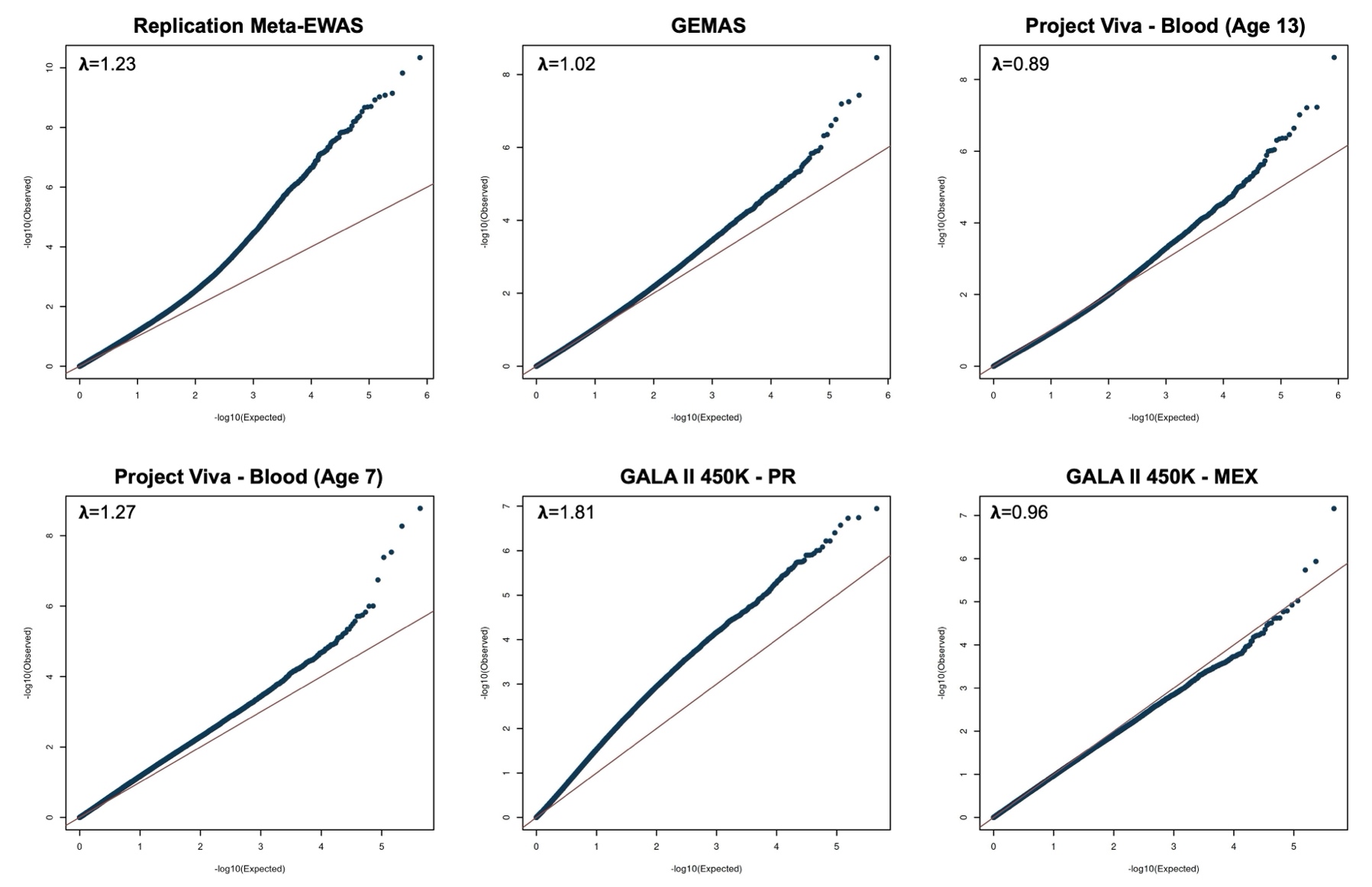

**Figure S6**. Quantile-Quantile (Q-Q) plots for the epigenome-wide meta-analysis and individual datasets in the replication phase. Uncorrected Q-Q plots and genomic inflation factors are shown. Large genomic inflation was detected and corrected only in GALA II 450K – PR using the BACON Bayesian method before the meta-analysis (λ_uncorrected_: 1.81, λ_corrected_: 1.15).

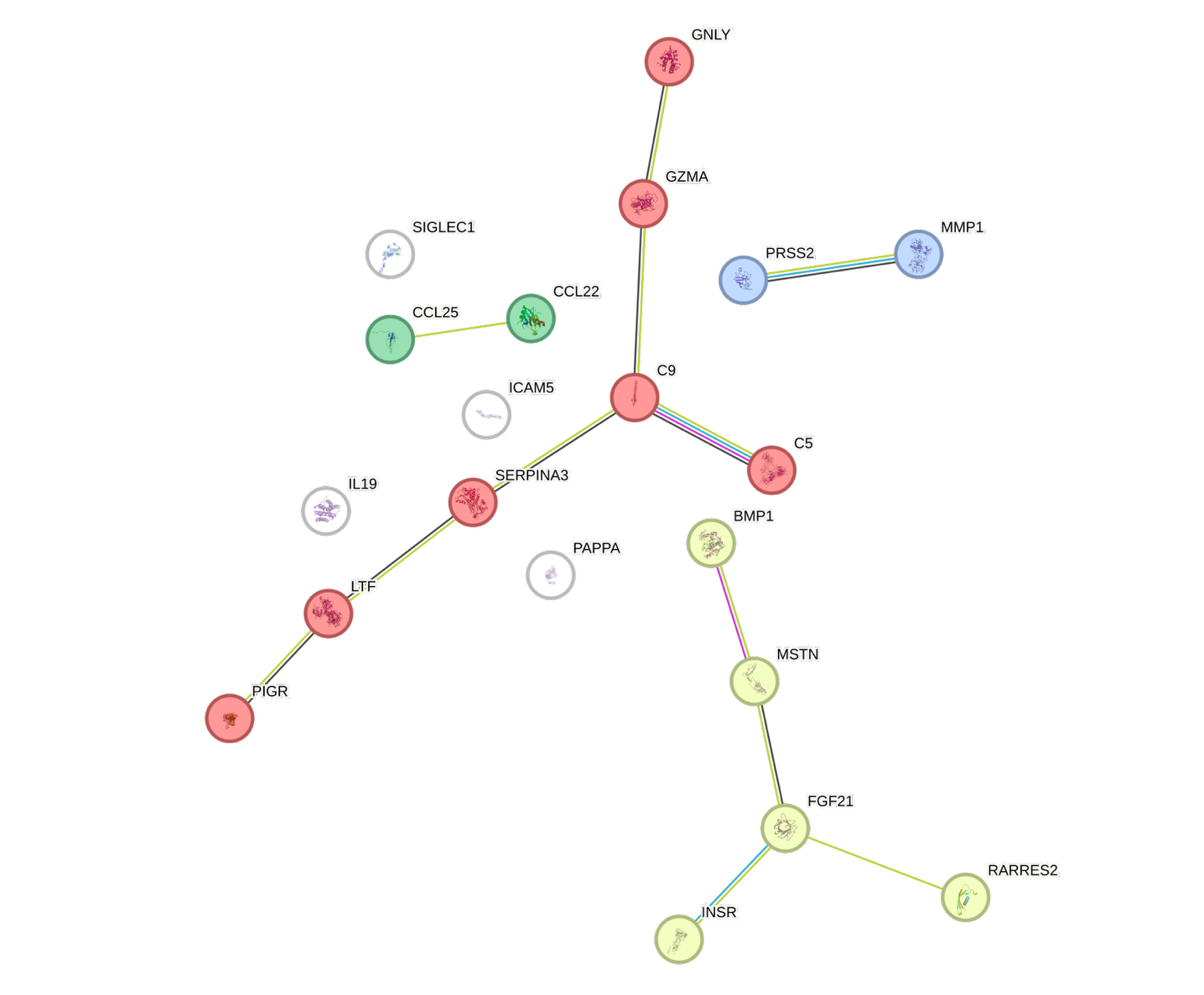

**Figure S7**. Protein-Protein functional and physical interaction (PPI) network based on DNAm-predicted plasma proteins associated with asthma exacerbations. Nodes represent the proteins and edges the PPI (based on a medium confidence score >0.4). PPI included known interactions from curated databases (cyan) and experimentally determined (pink), predicted interactions are based on gene neighborhood (green), gene fusions (red), and gene co-occurrence (dark blue), and associations related to text mining (yellow), co-expression (black), and protein homology (grey). Proteins were clustered in four groups using the k-means clustering method (red, yellow, green, and blue). Primary pathways for each cluster are cytolysis (red), mixed pathways including adiponectin binding and resistin (yellow), and activation of matrix metalloproteinases (blue). No primary description was provided for the green cluster.

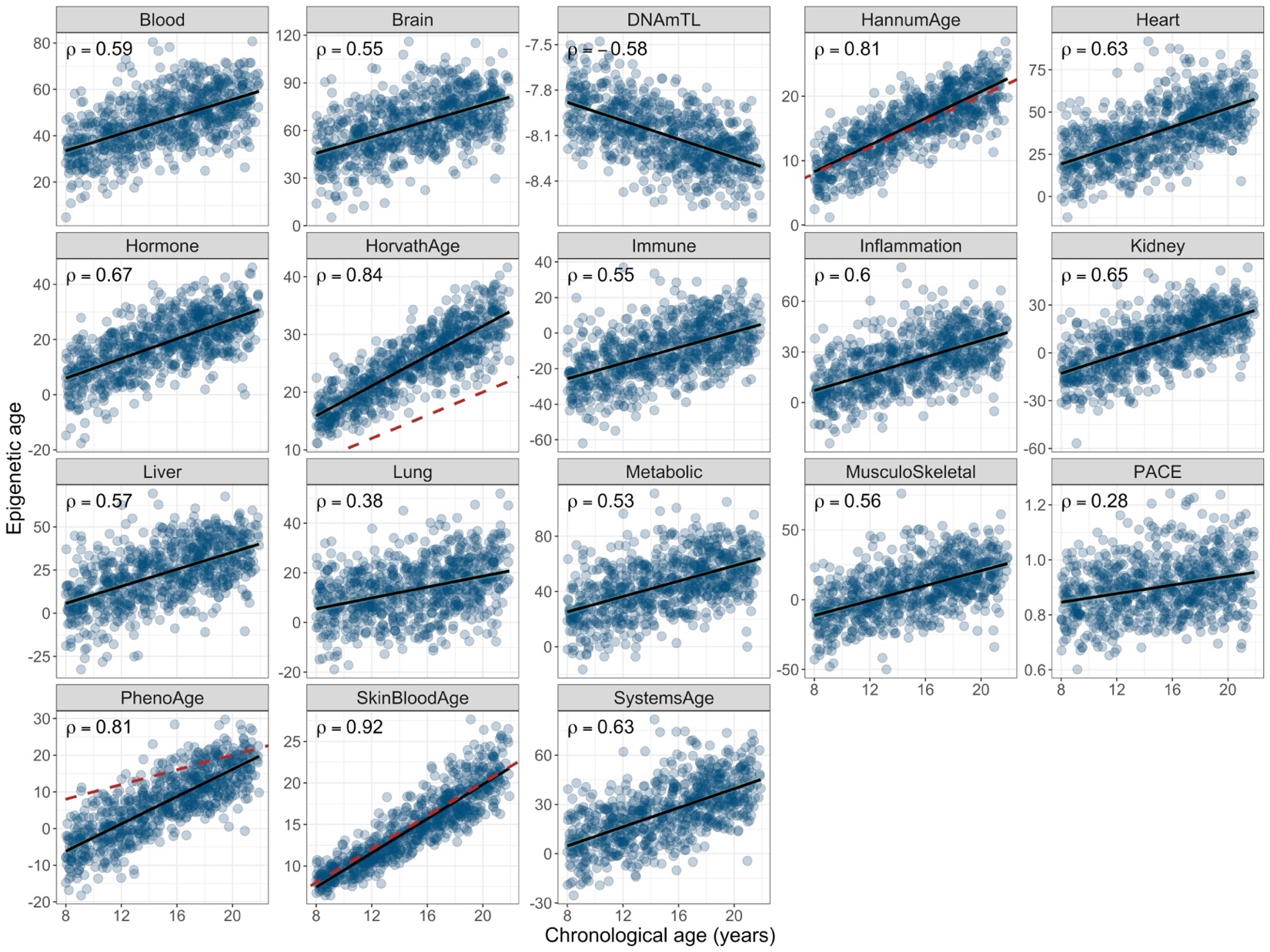

**Figure S8.** Panel of scatter plot showing the correlation between chronological age in years (x-axis) and epigenetic aging clocks (y-axis) in African Americans from SAGE. The black line shows the fitted regression line, with the 95% confidence band shown as a green shaded area. For clocks calibrated in years, the red dashed line indicates the 1:1 line where epigenetic age equals chronological age. The Pearson correlation coefficient is shown.

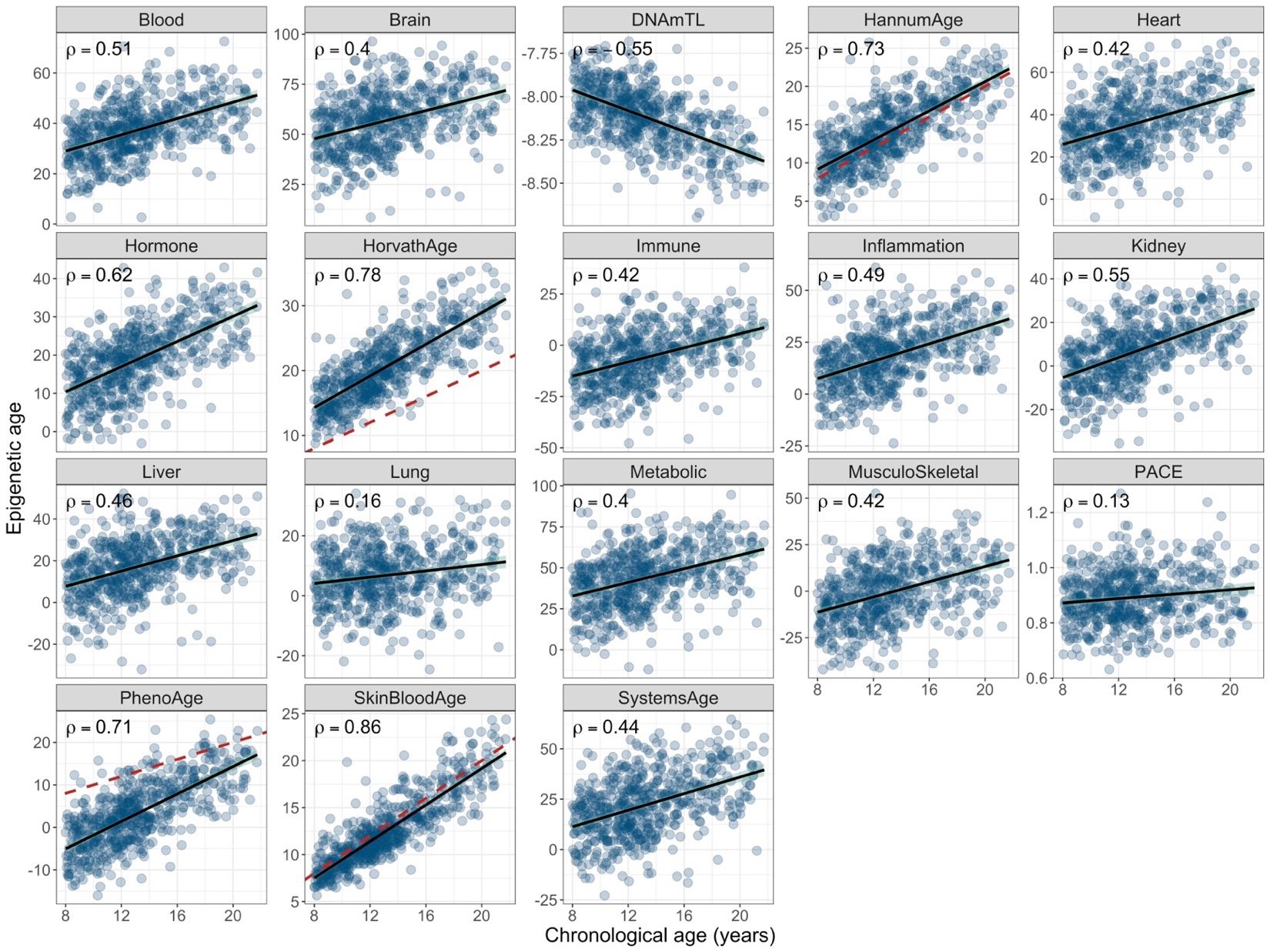

**Figure S9.** Panel of scatter plot showing the correlation between chronological age in years (x-axis) and epigenetic aging clocks (y-axis) in Puerto Ricans from GALA II. The black line shows the fitted regression line, with the 95% confidence band shown as a green shaded area. For clocks calibrated in years, the red dashed line indicates the 1:1 line where epigenetic age equals chronological age. The Pearson correlation coefficient is shown.

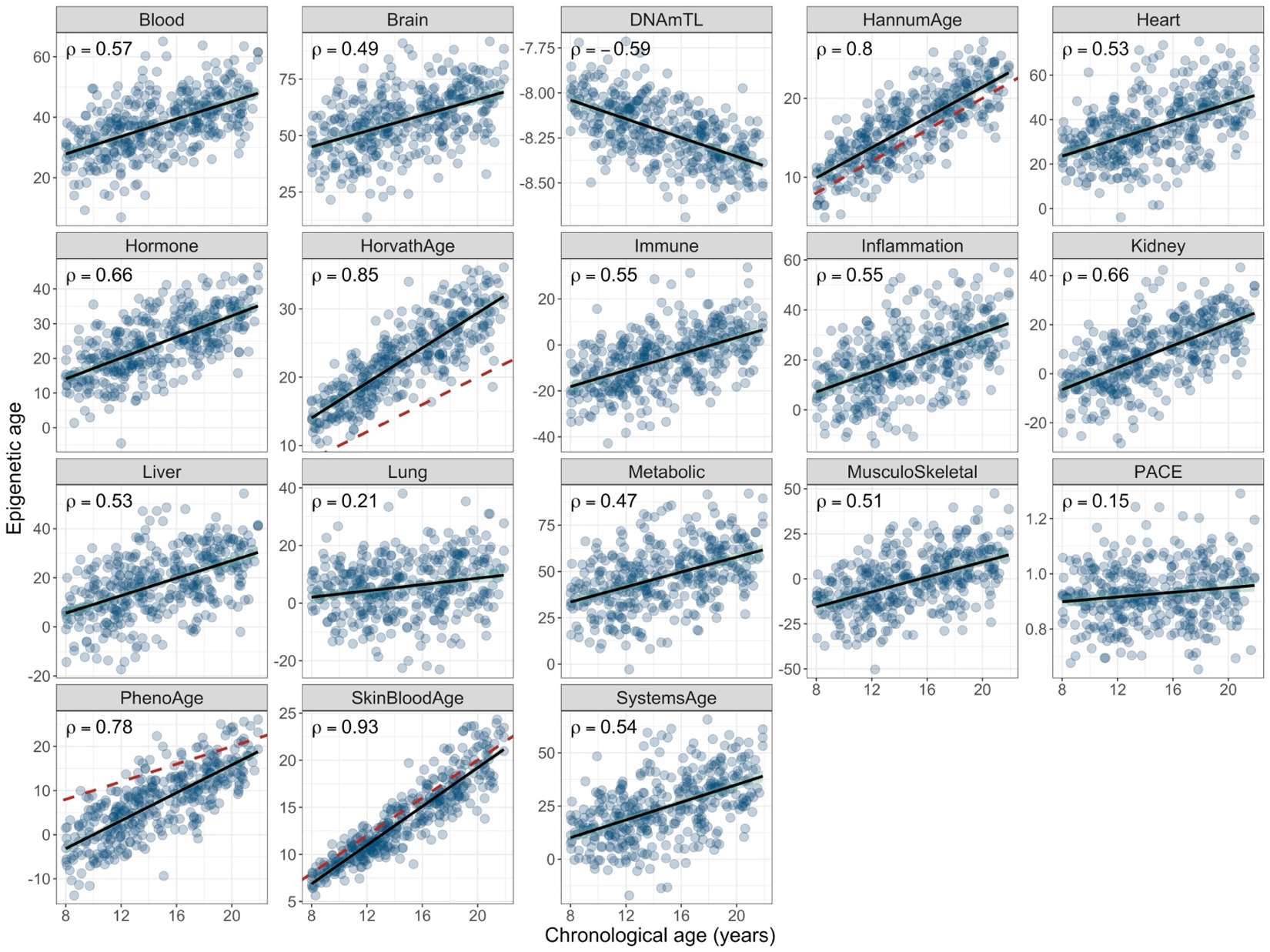

**Figure S10**. Panel of scatter plot showing the correlation between chronological age in years (x-axis) and epigenetic aging clocks (y-axis) in Mexican Americans from GALA II. The black line shows the fitted regression line, with the 95% confidence band shown as a green shaded area. For clocks calibrated in years, the red dashed line indicates the 1:1 line where epigenetic age equals chronological age. The Pearson correlation coefficient is shown.
